# Genetic Analysis of Primary Aldosteronism Bridging Disease Subtypes and the Phenotypic Continuum

**DOI:** 10.64898/2026.08.16.26359407

**Authors:** Daiki Tomidokoro, Fumihiko Takeuchi, Yuya Tsurutani, Yuta Tezuka, Masanori Murakami, Masahiro Nakatochi, Yuto Yamazaki, Yoshikiyo Ono, Takashi Suzuki, Ryoko Ishii, Mitsuhiro Yokota, Ken Yamamoto, Sahoko Ichihara, Hironobu Sasano, Akiyo Tanabe, Masakatsu Sone, Tetsuya Yamada, Fumitoshi Satoh, Tetsuo Nishikawa, Yukio Hiroi, Norihiro Kato

**Author notes:** Corresponding author Norihiro Kato, 1-21-1 Toyama, Shinjuku-ku, Tokyo, 162-8655, JAPAN.

## Abstract

Primary aldosteronism (PA) is a common cause of secondary hypertension. To investigate its genetic basis, we perform a trans-ancestry genome-wide association study (GWAS) meta-analysis, with subtype-specific analyses for aldosterone-producing adenoma (APA) and bilateral adrenal hyperplasia (BAH). Subsequently, we conduct genetic mediation analysis to partition PA effects on cardiovascular outcomes into blood pressure (BP)-mediated and BP-independent components. We further use a genetic risk score (GRS) to assess whether polygenic susceptibility to PA is associated with aldosterone-related traits in both population-based and PA case cohorts. We report 19 PA loci, including 13 new loci. While PA shares a broad polygenic framework across ancestries, subtype-specific heterogeneity exists, most notably at *TARID/TCF21*, which is preferentially associated with APA. A substantial proportion of the association between PA and cardiovascular disease is independent of systolic BP, particularly for heart failure and ischemic stroke. In population-based cohorts, a higher PA GRS is associated with higher systolic BP, lower serum potassium, and higher aldosterone levels, whereas in PA cases, particularly BAH, a higher GRS is linked to more severe aldosterone excess. Our results suggest that subclinical autonomous aldosterone excess exists along a continuous genetic spectrum across the population and that PA drives cardiovascular disease through substantial BP-independent pathways.

## Introduction

Primary aldosteronism (PA), estimated to affect 5–20% of patients with hypertension, is the most common and modifiable form of secondary hypertension. It is characterized by autonomous aldosterone secretion, leading to hypertension and/or hypokalemia, and an increased risk of stroke and myocardial infarction. Most PA cases are classified as unilateral aldosterone-producing adenoma (APA) or bilateral adrenal hyperplasia (BAH), also known as idiopathic hyperaldosteronism (IHA). Aldosterone production normally occurs in the zona glomerulosa (zG)—the outermost layer of the adrenal cortex—and abnormal aggregates of these cells, termed aldosterone-producing cell clusters (APCCs)/aldosterone-producing micronodules (APMs), are believed to constitute the common pathological basis of APA and BAH^1^.

PA occurs in familial and sporadic forms, with familial cases accounting for only about 5% of all PA. Four familial forms of PA (FH-I through FH-IV) have been recognized, each with distinct clinical manifestations and genetic causes^2^. In sporadic forms of PA, advances in somatic variant sequencing have enabled the identification of driver mutations in more than 90% of APA patients^3^. Despite arising in different genetic contexts, the affected genes converge on dysregulated aldosterone biosynthesis, involving genes that encode potassium channels (*KCNJ5*), calcium channels (*CACNA1D*, *CACNA1H*), chloride channels (*CLCN2*), ATPases (*ATP1A1*, *ATP2B3*), and mediators of the Wnt/β-catenin pathway (*CTNNB1*, *WNT2B*). Beyond rare and somatic variants, recent genome-wide association studies (GWAS) of PA have suggested that common germline variation also contributes to disease risk^4,5^.

Despite these pathophysiological insights, important knowledge gaps remain. Although the genetic architecture of PA is clearly multifactorial, the number of established susceptibility loci remains limited, and differences among PA subtypes and populations have not yet been fully explored. In addition to this lack of genetic data, the broader clinical implications of PA require clarification. Specifically, it remains unclear whether the increased cardiovascular risk in PA patients is primarily mediated by hypertension caused by aldosterone excess or whether it reflects aldosterone effects independent of blood pressure (BP)^6,7^. From a diagnostic perspective, conventional screening protocols based on plasma aldosterone and renin levels may fail to detect mild PA (i.e., autonomous aldosterone hyperproduction), which results in at-risk individuals being undetected. Furthermore, it remains unclear whether this mild phenotype forms a true pathophysiological continuum with overt PA.

To bridge this knowledge gap, we conducted a PA GWAS in Japanese individuals, followed by a trans-ancestry PA GWAS meta-analysis, to identify genetic characteristics specific to each subtype at the individual-locus level. In addition, we used genome-wide cohort data from large populations and publicly available GWAS summary statistics (Supplementary Table 1) to quantitatively assess the genetic relationships between PA and related traits, including systolic blood pressure (SBP), serum potassium, and cardiovascular disease (CVD). The results of this genetic study shed light on the broader clinical implications of PA and will help improve risk stratification and management strategies.

## Results

### PA GWAS in a Japanese Cohort

We conducted a PA GWAS in 670 cases from the Japanese PA Study (JPS) cohort and 25,998 non-hypertensive controls from BioBank Japan (BBJ)^8,9^ (Supplementary Table 2). Three loci reached genome-wide significance (*P* < 5.0 × 10^−8^). Two of these mapped to known PA-associated regions at 1p13 near *WNT2B* and 7p15 near *HOTTIP*, while the third represented a novel signal at 5q23 near *FBN2* (rs6595829, odds ratio [OR] = 1.42, 95% confidence interval [CI] = 1.26–1.59, *P* = 2.9 × 10^−8^) (Supplementary Fig. 1a, Supplementary Table 3).

### Trans-Ancestry PA GWAS Meta-Analysis

A trans-ancestry meta-analysis of PA GWAS was performed by combining Japanese results with publicly available European (Supplementary Fig. 1b) and African (Supplementary Fig. 1c) datasets. The combined datasets encompassed 3,706 cases (2,360 European, 670 Japanese, and 676 African) and 1,497,665 controls (Supplementary Table 4, Figure 1). The inverse-variance weighted (IVW) meta-analysis identified 18 significant loci (Supplementary Table 5). Subsequent trans-ancestry meta-regression using MR-MEGA^10^ identified 14 loci that largely overlapped with the IVW findings but included one additional locus near *CLRN1* (Supplementary Table 6). Together, these approaches yielded 19 PA loci, including 13 new discoveries (Table 1, Figure 1). Evaluation of the inter-ancestry genetic architecture revealed a strong genome-wide correlation (ρ_ge_ = 0.80, SE = 0.30) between European and Japanese cohorts. However, at the individual-locus level, significant inter-ancestral heterogeneity was observed at the *CLRN1* and *RXFP2* loci (*P* < 2.6 × 10^−3^) (Supplementary Data 1, Supplementary Fig. 2).

**Figure 1.**
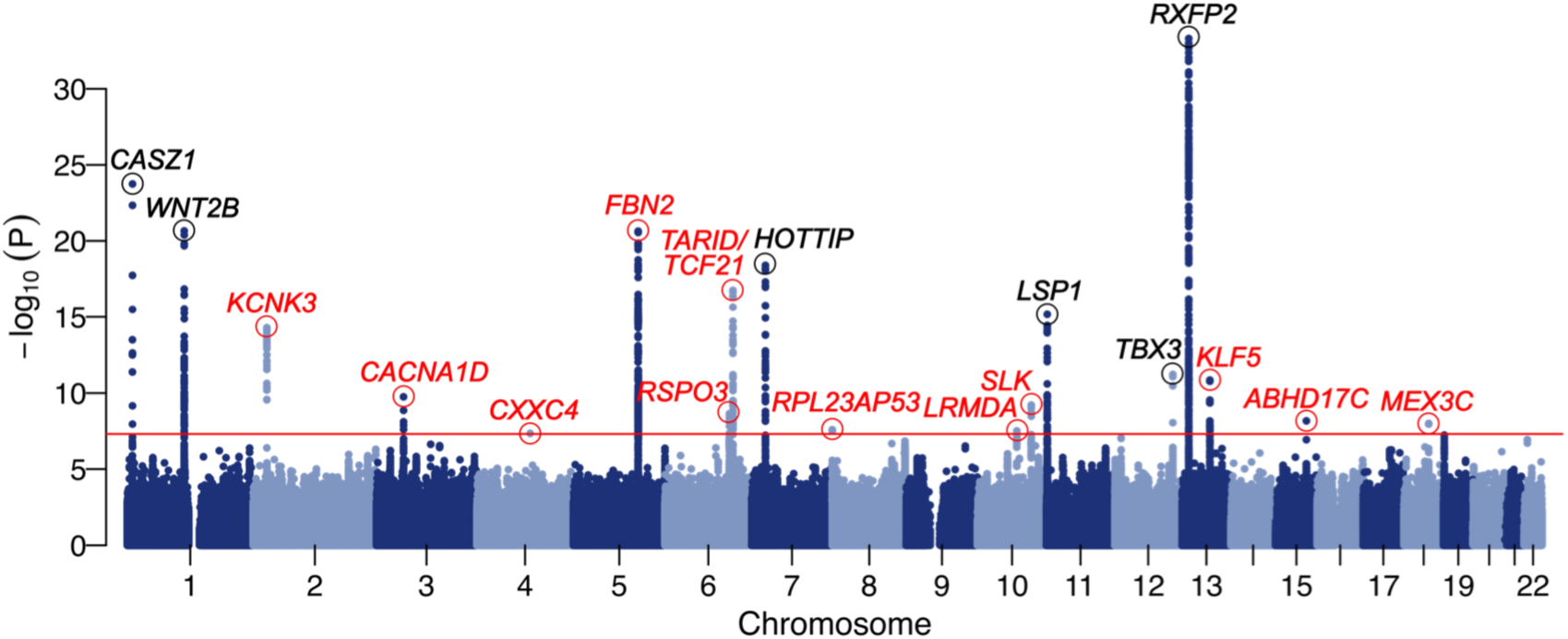
Trans-ancestry PA GWAS. A Manhattan plot showing genome-wide significant associations. The red horizontal line denotes genome-wide significance (*P* < 5.0 × 10^−8^). Red text highlights new loci, while black text indicates previously reported loci. The dots enclosed in circles are lead SNPs at each locus.

**Table 1.**
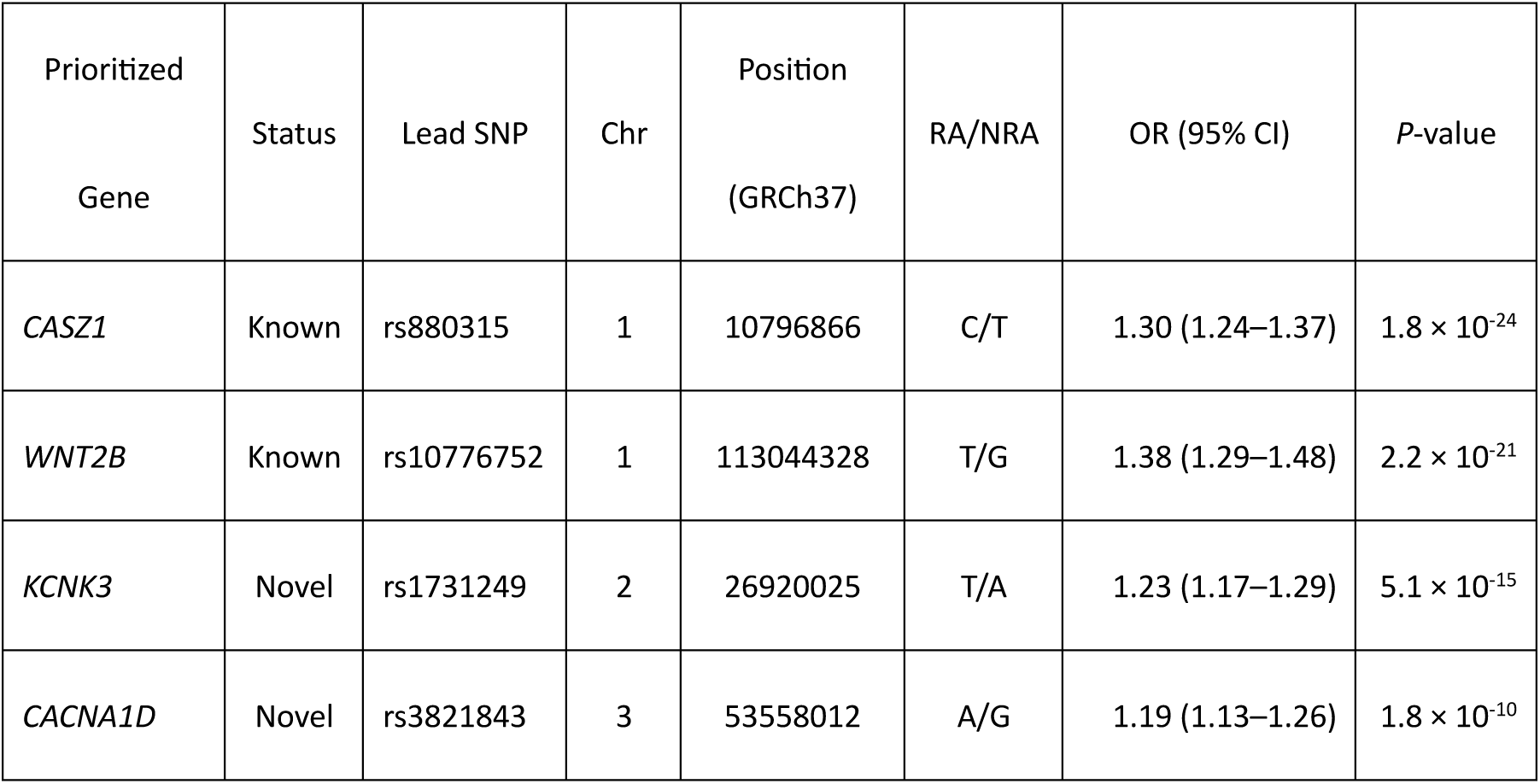
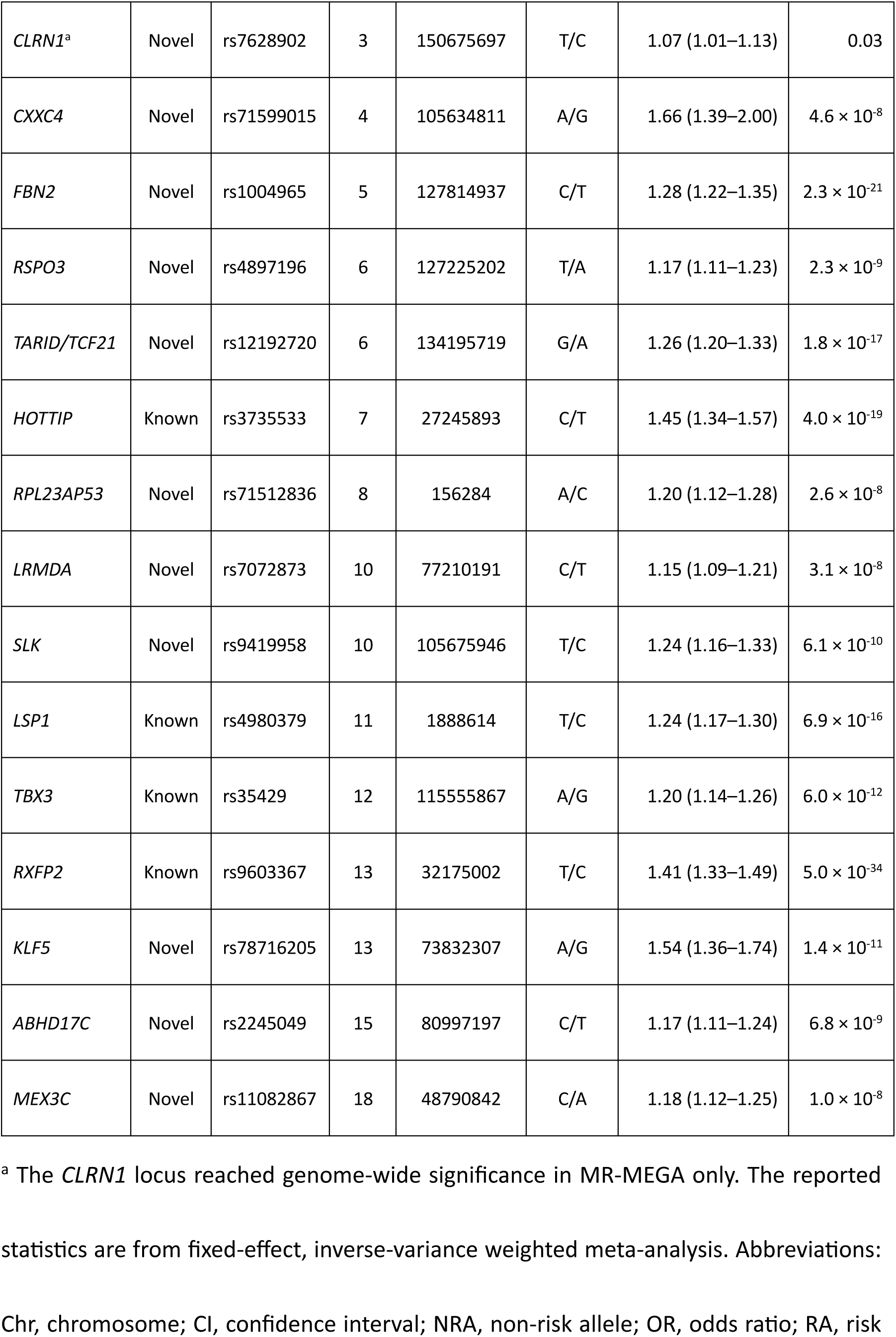
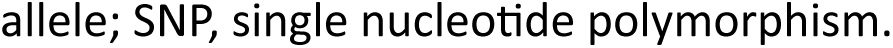
Significant loci identified in the trans-ancestry PA GWAS meta-analysis.

### Fine-Mapping and Functional Annotation

Trans-ancestry fine-mapping of the 19 PA loci identified 20 credible sets across 17 loci, with a median size of 4 variants (range 1–25 variants per credible set; Supplementary Data 2). Eighty-two (59.9%) of the 137 credible-set variants had strong adrenal regulatory annotations (RegulomeDB^11^ Rank ≤ 2 or FORGEdb^12^ Score ≥ 8), with enrichment in enhancers, promoter-proximal regions, and open chromatin (Supplementary Table 7). Lead single nucleotide polymorphisms (SNPs) at eight loci showed significant expression quantitative trait loci (eQTL) effects on 13 genes, and two splicing quantitative trait loci (sQTL) signals affecting distinct *LSP1* splice junctions were detected at one locus (Supplementary Data 3 and 4, Supplementary Fig. 3). PA association signals colocalized with the regulation of mRNA expression of *KCNK3*, *LRMDA*, *ABHD17C*, and *MEX3C*, as well as the splicing of *LSP1* (PP.H4 > 0.8; Supplementary Tables 8 and 9). In particular, multiple lines of evidence supported the regulatory mechanism for the fine-mapped variant at 15q25 to influence *ABHD17C* expression (Supplementary Fig. 4). We also detected a long-range chromatin interaction between the 10q24 locus and *SLK* (false discovery rate [FDR] = 1.0 × 10^−6^, Table 1).

### Subtype-Stratified PA GWAS

Within the Japanese cohort, subtype-stratified PA GWAS (301 APA cases, 294 BAH cases) identified an APA locus near *TARID/TCF21* and a BAH locus at *HOTTIP*, both consistent with the primary PA meta-analysis (Supplementary Table 10, Supplementary Fig. 5).

Meta-analyses combining the Japanese JPS cohort and the French COMETE (COrtico-et MEdullo-surrénale, les Tumeurs Endocrines) network^5^ cohorts (a total of 622 APA cases and 534 BAH cases) revealed several associations specific to each subtype. The APA meta-analysis confirmed the association near *TARID/TCF21* on chromosome 6 (lead SNP rs36125724, OR = 1.59, 95% CI = 1.41–1.79, *P* = 2.7 × 10^−14^) (Supplementary Fig. 6a, Supplementary Table 11). The BAH meta-analysis also strengthened the association near *HOTTIP* on chromosome 7 (lead SNP rs1859168, OR = 1.60, 95% CI = 1.36–1.87, *P* = 8.6 × 10^−9^), and detected a second significant locus near *CASZ1* on chromosome 1 (lead SNP rs880315, OR = 1.51, 95% CI = 1.31–1.73, *P* = 5.5 × 10^−9^) (Supplementary Fig. 6b, Supplementary Table 11).

Since there was significant overlap in susceptibility loci between the analyses of the overall PA and those stratified by subtype, we compared the effect estimates across subtypes for all PA lead SNPs (Figure 2a, Supplementary Table 12). Although the effects at each locus tended to correlate between the two subtypes, at the *TARID/TCF21* locus (a locus with ≥3 sentinel SNPs that exhibits allelic heterogeneity and complex genetic structure; Supplementary Fig. 7 and 8), significant heterogeneity between subtypes was observed for these sentinel SNPs (e.g., rs12192720, a trans-ancestry lead SNP with *P_het_* = 2.1 × 10^−4^) with a stronger effect detectable in APA (Figure 2a). Regional association plots showed a clear signal in APA but not in BAH (Figure 2b). In the GTEx^13^ adrenal data, the risk-increasing C allele at the APA lead SNP (rs36125724; Figure 2c) was associated with reduced *TCF21* mRNA expression (*P* = 2.7 × 10^−9^; Figure 2d), which could lead to tumorigenesis in APA.

**Figure 2.**
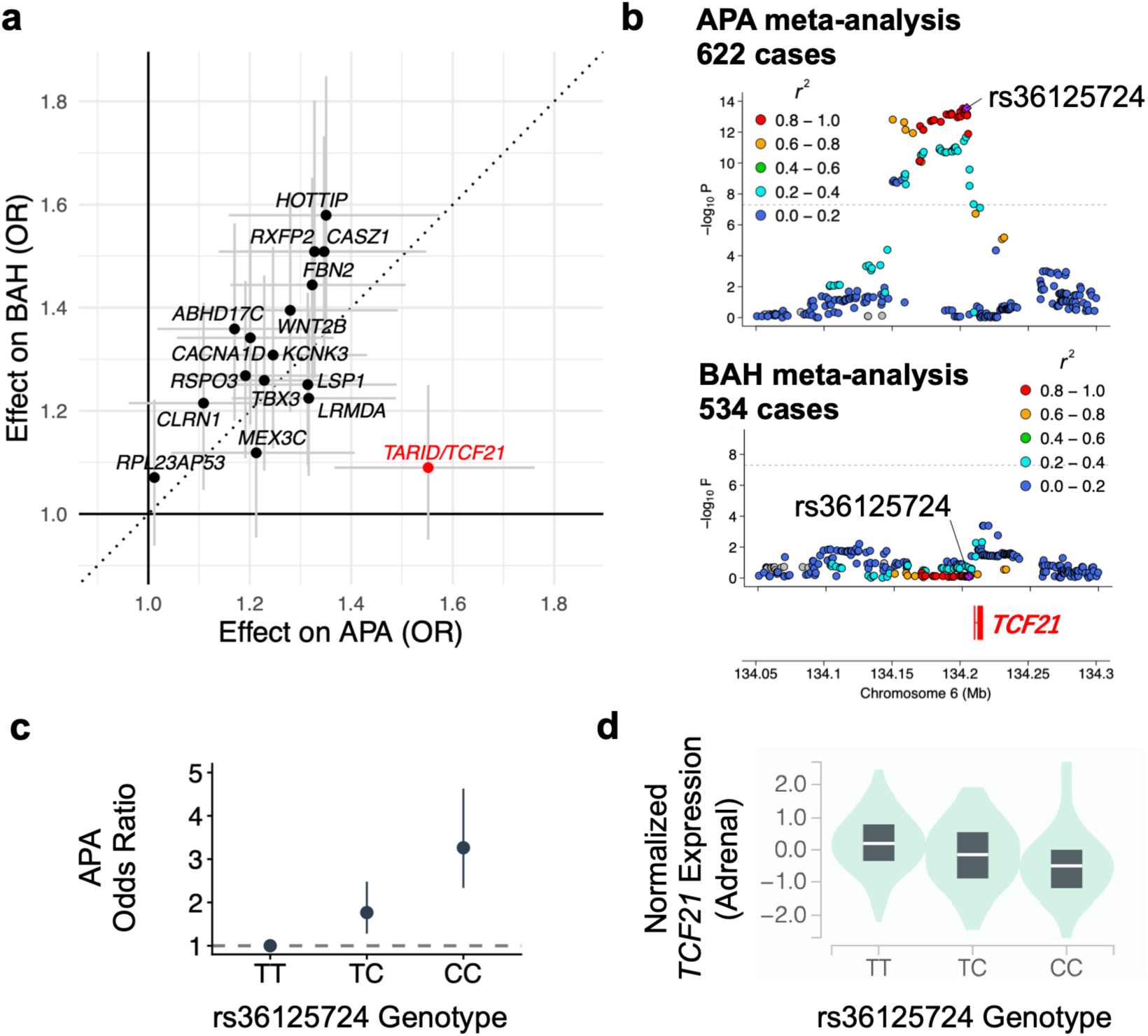
Subtype-specific genetic architecture of PA at the *TARID/TCF21* locus. **(a)** A scatter plot comparing the odds ratios (OR) for APA versus BAH at the lead SNPs detected in the PA GWAS meta-analysis. **(b)** Regional Manhattan plots for APA meta-analysis (top) and BAH meta-analysis (bottom). **(c)** Association of rs36125724 genotype with APA risk in the Japanese dataset. OR (points) and 95% confidence intervals (error bars) are shown for each genotype relative to the TT genotype. **(d)** A violin plot illustrating the significant association between the rs36125724 genotype and *TCF21* mRNA expression levels in the adrenal glands in the GTEx dataset.

### Comparison of Genetic and Biochemical Measures Between PA Subtypes

To investigate the cumulative effects of susceptibility loci, we constructed genetic risk scores (GRSs) for two ancestral groups (Europeans and Japanese) using a specific subset of lead SNPs (Supplementary Table 13). As mentioned earlier, because the *TARID/TCF21* locus showed a distinctive pattern of effect sizes across the two subtypes (Figure 2a and Supplementary Table 12), we used two genetic measures—the rs36125724 genotype and the modified GRS excluding this locus—when comparing the two subtypes within JPS PA cases (301 APA and 294 BAH cases).

We found significant differences between the subtypes in the following measures: 24-hour urinary aldosterone excretion (U-Ald) was higher in the APA group (*P* < 2.2 × 10^−16^), while serum potassium levels and the modified GRS were both lower in APA than in BAH (*P* < 2.2 × 10^−16^ and *P* = 0.005, respectively; Supplementary Fig. 9 and Supplementary Table 2). Focusing on the modified GRS (i.e., cumulative effect excluding *TARID*/*TCF21*), 24-hour urinary aldosterone excretion increased significantly in BAH (adjusted *P* = 0.0036) as the GRS increased. While APA demonstrated a similar trend, the slope of association was steeper for BAH than for APA (Supplementary Fig. 10 and Supplementary Table 14). On the other hand, when focusing on the rs36125724 genotype, no significant positive correlation was observed between the number of rs36125724 risk alleles associated with tumorigenesis and 24-hour urinary aldosterone excretion in either disease subtype (Supplementary Fig. 11a).

### BP-Independent Effects on CVD

LD score regression (LDSC)^14^ analyses revealed significant genetic correlation between PA and major CVD outcomes. In Europeans, PA showed significant genetic correlations with coronary artery disease (CAD) (*r*_g_ = 0.23 ± 0.06), atrial fibrillation (AF) (*r*_g_ = 0.21 ± 0.06), heart failure (HF) (*r*_g_ = 0.38 ± 0.06), and ischemic stroke (IS) (*r*_g_ = 0.44 ± 0.07), and the strength of these correlations was comparable to that observed for SBP (Figure 3a). A similar trend was observed in Japanese individuals (Supplementary Table 15).

**Figure 3.**
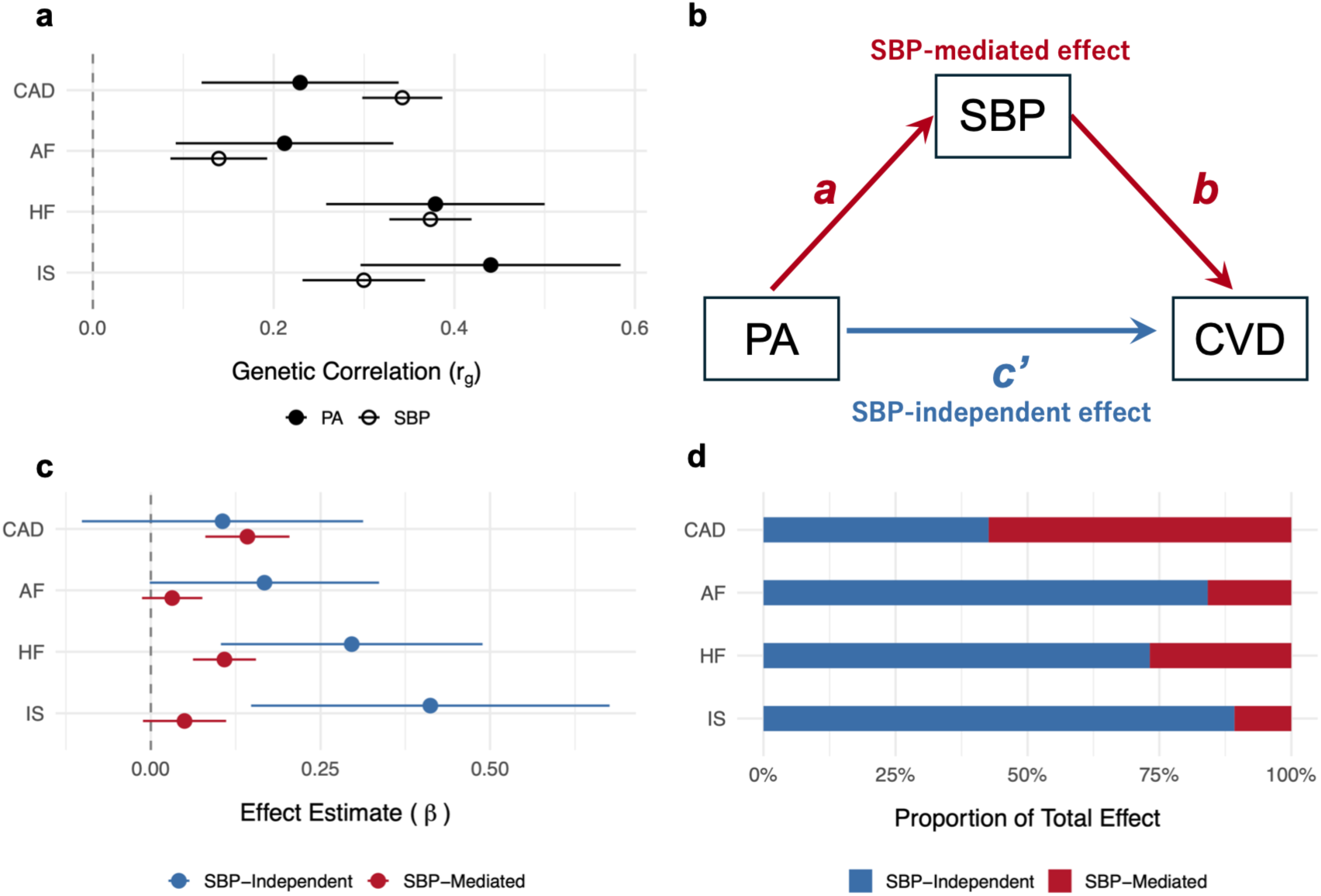
BP-dependent and -independent genetic effects of PA on CVD. **(a**) A forest plot showing the genetic correlations between PA and four CVDs, as well as the corresponding correlations with SBP. **(b**) A schematic diagram of the Genomic SEM model used to partition the total genetic effect of PA on CVD into an SBP-mediated pathway (paths *a* and *b*) and an SBP-independent pathway (*c’*). (**c**) A forest plot showing SBP-independent effect (*c’*; blue) and SBP-mediated effect (*a* × *b*; red) estimates of PA on each CVD. Error bars represent 95% CI. **(d)** Stacked bar chart illustrating the relative contributions of the SBP-independent (blue) and SBP-mediated (red) pathways.

In genetic mediation analyses among European individuals (Figure 3b), PA exhibited significant (*P* < 0.003) SBP-independent effects on HF (*c’* = 0.30), and IS (*c’* = 0.41) (Figure 3c, Supplementary Table 16). SBP-independent effects were also observed for CAD (*c’* = 0.11) and AF (*c’* = 0.17), although these did not reach statistical significance. The proportion of the total genetic effect acting independently of SBP was estimated to be 42.6% for CAD, 84.2% for AF, 73.2% for HF, and 89.2% for IS (Figure 3d). In analyses of Japanese individuals, point estimates of a similar magnitude were obtained for these non-mediated effects (Supplementary Table 16). In the sensitivity analysis, the non-mediated proportions for HF, IS, and AF in the European cohort remained stable even after adjustment for metabolic covariates (Supplementary Table 17).

### Genetic Overlap of PA with Blood Pressure and Serum Potassium

Given that individuals with higher BP may have a stronger predisposition to PA, this study employed a case-control study design for the Japanese PA GWAS, using non-hypertensive individuals as the control group. Among the 56 top SBP-associated loci in East Asians^15^, the alleles associated with higher BP were also associated with an increased risk of PA at the nominal significance level (*P* < 0.05) in 18 loci (32.1%). However, at 9 loci (16.1%), the direction of the association with PA was reversed (Supplementary Fig. 12, Supplementary Data 5). These findings were consistent with a previous report^4^. Notably, loci with large effects on SBP but weak association with PA (e.g., *ALDH2* and *FGF5*) have been implicated in BP regulation through mechanisms largely unrelated to PA^16,17^.

In turn, among the 18 PA loci (excluding the *CLRN1* locus, for which no genetic association was observed in Europeans), 15 loci showed genome-wide significant associations with both SBP and serum potassium in Europeans. At all loci with available data (18 for serum potassium and 17 for SBP), the risk allele for PA was associated with an increase in SBP and a decrease in serum potassium levels (Figure 4a, Supplementary Table 18).

**Figure 4.**
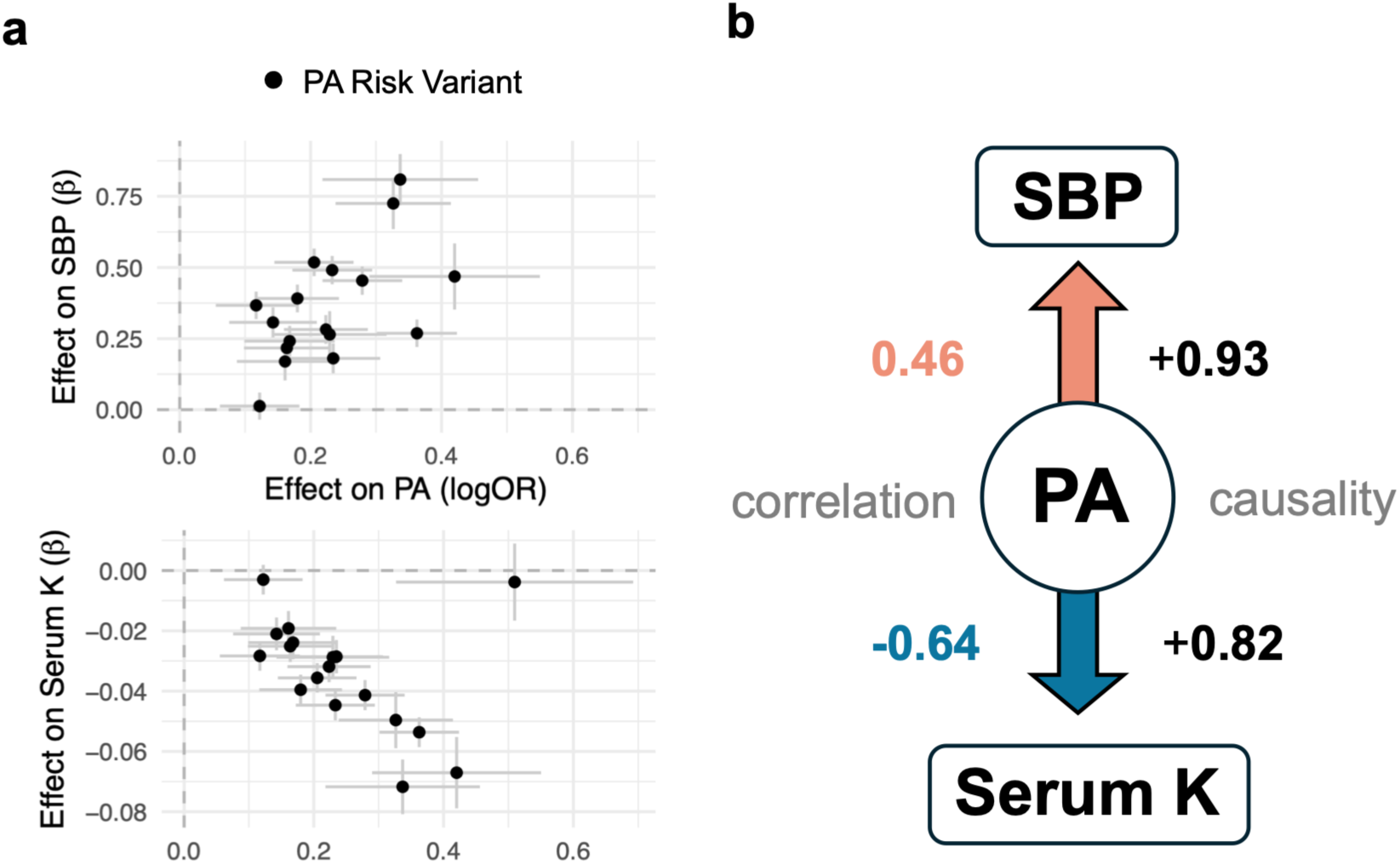
Locus-level and genome-wide relationships of PA susceptibility with SBP and serum potassium. **(a)** Scatter plots showing the relationship between the effect size (log_10_OR) of the PA-lead variant and its respective effects (betas) on SBP (top) and serum potassium (serum K; bottom) in European cohorts. The *CXXC4* locus is excluded from the SBP plot due to missing data. **(b)** A schematic diagram illustrating genome-wide genetic correlations and causal pathways from PA to SBP and from PA to serum potassium.

### Causal Modeling of Aldosterone-Related Traits

Across the genome, PA showed a strong positive genetic correlation with SBP (*r*_g_ = 0.46 ± 0.06, *P* = 2.3 × 10^−13^) and a strong negative correlation with serum potassium (*r*_g_ = −0.64 ± 0.08, *P* = 7.7 × 10^−15^) in Europeans (Figure 4b). Among Japanese individuals, similar correlations were observed for SBP (*r*_g_ = 0.54 ± 0.11, *P* = 9.0 × 10^−7^) and serum potassium (*r*_g_ = −0.19 ± 0.11, *P* = 0.07). Latent causal variable (LCV)^18^ modeling further confirmed the causal effects of PA on both SBP and serum potassium, with estimated proportions of genetic causality of +0.93 (SE = 0.06, *P* = 9.4 × 10^−32^) for SBP and +0.82 (SE = 0.14, *P* = 9.5 × 10^−7^) for serum potassium (Figure 4b).

### Population-Level Impact of PA Polygenic Risk

The distribution of PA GRS was compared among Japanese PA cases (JPS cohort; *n* = 670), the entire BBJ cohort (*n* = 51,071), and the Kita-Nagoya Genomic Epidemiology (KING) study^19^ cohort (*n* = 2,453) (Figure 5a). As expected for a polygenic trait, the GRS followed a normal distribution in all cohorts. The GRS in the PA cases was significantly higher than in both BBJ (+0.59 SD, *P* < 0.001) and KING (+0.62 SD, *P* < 0.001) cohorts, but there was no significant difference between BBJ and KING cohorts (*P* = 0.32).

**Figure 5.**
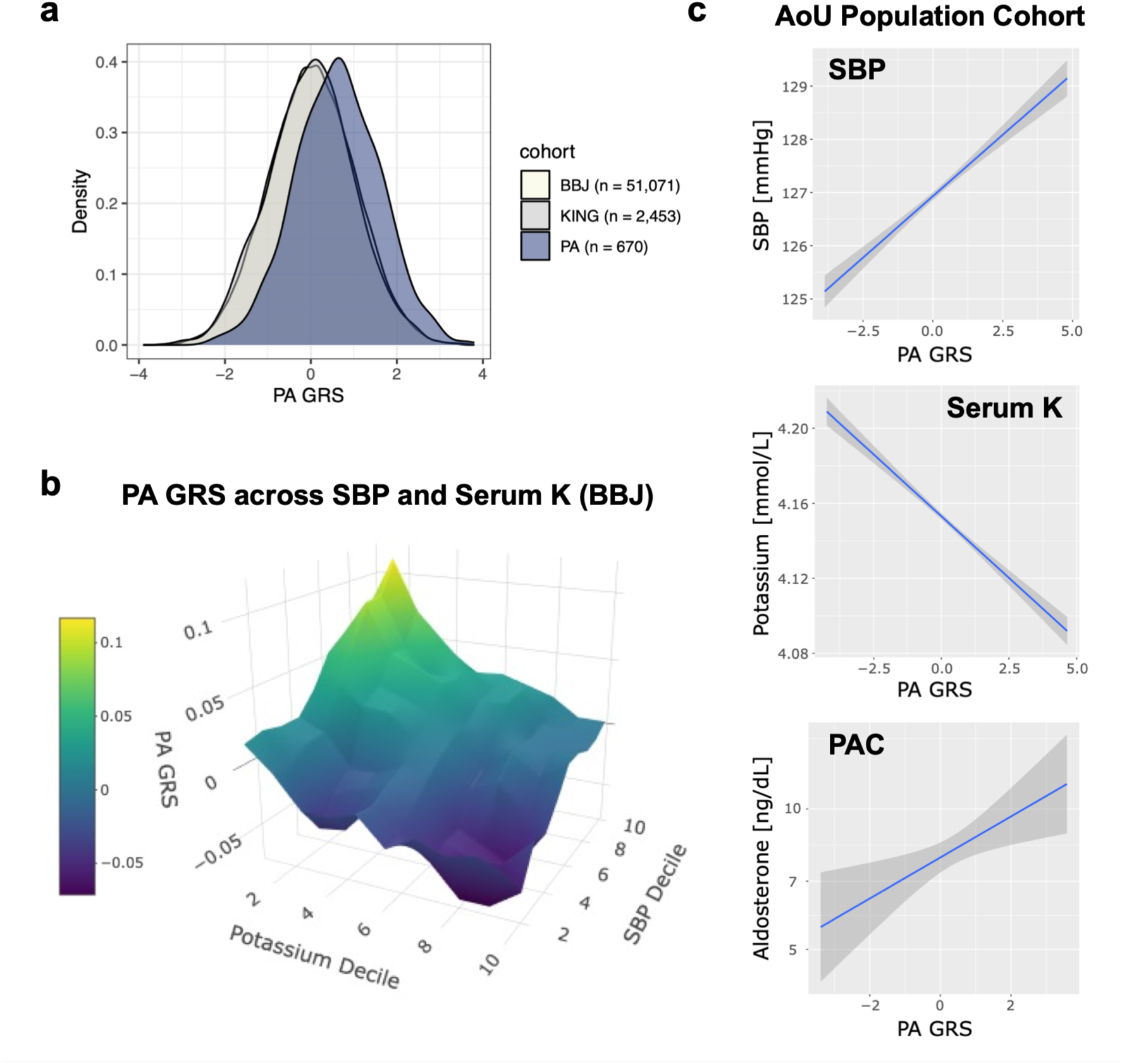
Distribution of PA GRS and its association with PA-related traits across multiple cohorts. **(a)** Density plots showing the distribution of PA GRS in Japanese PA cases compared to two population-based cohorts: BBJ and KING (overall difference in GRS between PA cases and two cohorts: *P* < 2 × 10^−16^). **(b)** A 3D surface plot of the mean PA GRS mapped by deciles of SBP and serum potassium levels in BBJ, with spatial smoothing applied using a 3×3 mean filter. **(c)** Linear regression plots displaying associations of European PA GRS with SBP (top), serum potassium levels (middle), and log-transformed plasma aldosterone concentration (bottom) in the European subset of the AoU cohort.

In the BBJ cohort, PA GRS exhibited a continuous gradient across the entire range of SBP and serum potassium levels, peaking among individuals in the decile group with the highest SBP and the lowest serum potassium levels (Figure 5b). Linear regression analyses confirmed these findings (Supplementary Table 19). Specifically, higher PA GRS was significantly associated with increased SBP (*P* = 1.1 × 10^−10^) and lower serum potassium levels (*P* < 2.2 × 10^−16^). Similarly, significant associations were observed between PA GRS and SBP in the Japanese KING cohort, and between PA GRS and both traits in the All of Us (AoU)^20^ cohort (*P* < 0.02 in both instances; Figure 5c, Supplementary Table 19). Furthermore, in the AoU cohort, a higher GRS was significantly associated with elevated plasma aldosterone concentration (PAC) (*P* = 0.005; Figure 5c).

## Discussion

In this multi-ancestry GWAS, we have identified 19 PA loci, including 13 new associations. This study is particularly notable for its novelty in the following three areas. First, while individual risk alleles generally exhibit a consistent direction across ancestries, the broader genetic architectures differ between subtypes: BAH is characterized by a markedly high polygenic burden, whereas genetic susceptibility to APA is strongly influenced by significant subtype-specific effects at the *TARID/TCF21* locus. Second, according to genetic mediation analysis, the association between PA and adverse cardiovascular outcomes is suggested to involve pathways that operate independently of BP regulation. Third, genetic predisposition to PA is associated with interindividual variability in aldosterone levels in both the general population and cases clinically diagnosed with BAH, and has downstream effects on BP and serum potassium levels.

Our GWAS meta-analysis not only replicates previously reported PA-associated loci (e.g., *CASZ1, WNT2B*, *HOTTIP*, and *LSP1*) but also identifies new loci (as shown in Table 1), thereby deepening our understanding of the pathways involved in this disease. These results suggest two principal mechanisms, consistent with previous findings—regulation of zG membrane potential by ion channels and Wnt signaling—as well as other pathways related to aldosterone biosynthesis and function (Supplementary Fig. 13). With regard to ion channels, genes such as *CACNA1D*, encoding a Ca²⁺ ion channel; *KCNK3*, encoding a TASK1 K⁺ leak channel; and *ABHD17C* (Supplementary Fig. 4), encoding a depalmitoylase implicated in the regulation of Ca²⁺-activated K⁺ channels^21^, were located near PA loci. With regard to Wnt signaling, genes encoding components of two branches of this pathway—namely, non-canonical Wnt/planar-cell-polarity signaling (*WNT2B*)^22^ and canonical Wnt/ *β* -catenin signaling (*RSPO3* and *CXXC4*)^23–26^—were also located near the PA locus. Some of these target genes (e.g., *CYP11B1/CYP11B2* and *CACNA1D*) play a pathogenic role not only in genetic variants associated with sporadic PA but also in somatic mutations in APA and/or germline mutations in familial PA^27^ (Supplementary Table 20). This supports the notion that the underlying genetic mechanisms converge on a shared set of pathways. Next, we provide further explanation regarding the three areas of note in this study.

In the subtype-stratified GWAS meta-analysis, we demonstrate that the *TARID/TCF21* locus exhibits significant heterogeneity and preferentially affects APA. Although limited statistical power precludes a formal colocalization analysis, the locus overlaps with the *TCF21* eQTL (at rs36125724), and the APA risk allele is associated with reduced expression of that gene (Fig. 2d). TCF21 is a key regulatory protein whose expression is significantly reduced in adrenocortical carcinomas (ACCs)^28^. This protein functions as a tumor suppressor by negatively regulating steroidogenic factor 1 (SF-1/NR5A1), the primary protein responsible for adrenal cell proliferation and the transcription of genes (such as Steroidogenic Acute Regulatory protein [StAR]) necessary for steroidogenesis^28^. Given that elevated *SF-1* expression promotes the formation of adrenocortical tumors in murine models^29^ and that *SF-1* expression is upregulated in human APA^30^, the reduction in *TCF21* expression caused by risk alleles may weaken the repression of *SF-1*, thereby creating a state that permits clonal adrenocortical proliferation. While most ACCs are hormonally inactive or produce excessive cortisol, a very small proportion (about 2.5%) of ACCs predominantly secrete aldosterone^31^.

In light of these existing research findings, the following “two-step hit” hypothesis (Supplementary Fig. 11b) can be proposed regarding the findings of this study related to *TCF21*—specifically, that APA is associated with significantly higher aldosterone excretion than BAH (Supplementary Fig. 9a), yet no significant correlation was observed between the rs36125724 genotype (at *TCF21*) and aldosterone excretion in either disease subtype (Supplementary Fig. 11a). Although this remains purely hypothetical at this point, from the perspective of the progression from BAH to APCC/APM and then to APA, genetic factors other than the *TCF21* locus involved in aldosterone biosynthesis can influence all of these conditions. On the other hand, while risk alleles at the *TCF21* (“first hit”) locus increase tumorigenic potential (i.e., progression to APA via APCC/APM), the *TCF21* locus alone appears to have no noticeable effect on aldosterone biosynthesis. When somatic mutations (“second hit”) occur in APA-causing genes such as *KCNJ5*, tumor formation is promoted, and excessive aldosterone production manifests as APA; however, since the amount of aldosterone biosynthesized is strongly influenced by tumor size^32^, the impact of other genetic factors (e.g., the modified GRS excluding *TCF21*) becomes relatively smaller. Thus, the effect of germline variants on aldosterone production in APA becomes apparent when considered alongside somatic-level events, but it may not be as significant as that seen in BAH.

The significant BP-increasing effects observed at each PA locus (Figure 4a and Supplementary Fig. 12), as well as the strength of the genome-wide genetic correlation between PA and BP traits (Figure 4b), support the notion—as previously reported^4^—that PA makes a significant genetic contribution to hypertension. In addition, data from observational studies to date indicate that PA confers cardiovascular risk beyond that predicted by BP alone^7,33,34^. It remains difficult to determine whether this excess risk is due to the direct effects of aldosterone on cardiovascular tissues or is partly attributable to residual confounding factors. Recent Mendelian randomization studies have revealed a genetic association between PA and CVD (e.g., CAD, HF, and stroke)^35^; however, the effect of PA on CVD independent of BP has not been quantitatively assessed. The results of the genetic mediation analysis indicate that a large proportion of the genetic association between PA and CVD is independent of SBP (Figure 3). The proportion of SBP-independent effects exceeds 80% for both AF and IS, and is 73% for HF. Also, significant non-mediated effects (*c’*) are observed for HF and IS (Supplementary Table 17). The finding that AF and IS have a high proportion of SBP-independent effects is consistent with previous studies showing that these conditions exhibit the most disproportionate increases in disease risk in PA (pooled ORs of 3.52 and 2.58, respectively, compared with essential hypertension)^7^. In contrast, the small proportion of SBP-independent risk (43%) for CAD suggests that the coronary risk associated with PA is more strongly influenced by BP. Overall, these findings provide the first genomic estimate of the SBP-independent association between PA and CVD and support the potential for aldosterone-targeted therapies beyond BP management.

Observational studies suggest that renin-independent aldosterone secretion extends beyond cases of overt PA to encompass the entire BP continuum^36,37^. From a genetic perspective, although the heritability of aldosterone concentration is moderate^38^, no definitive genetic loci have been identified in previous GWAS of serum aldosterone^39^, and insights into its genetic basis remain limited. In this study, we show that polygenic susceptibility to PA is associated with dose-dependent effects on aldosterone levels and related phenotypes, even in individuals not diagnosed with PA (Figure 5b,c). Taken together, these results support a PA continuum model linking polygenic predisposition to aldosterone biology, and the PA GRS has the potential to quantify individual risk and serve as a guide for screening and early intervention.

In a recent preprint^40^, Fergus *et al.* reported a PA GWAS meta-analysis based exclusively on large-scale biobank data, identifying significant association signals overlapping with those in this study, and constructing a polygenic risk score that proved useful for distinguishing non-functional adrenal adenomas from PA. In contrast, our genetic study analyzed PA specifically from the perspectives of differences across disease subtypes (BAH vs APA), its clinical significance in the general population, and the pathogenic mechanisms underlying its cardiovascular complications.

Several limitations must be considered. First, from the perspective of the robustness of the trans-ancestry analysis, while the European dataset had sufficient statistical power, the non-European datasets had a relatively small sample size. Second, the dichotomy between APA and BAH has inherent limitations: from a biological perspective, it is an oversimplification (as evidenced by overlapping or transitional conditions such as APCCs/APMs^41,42^); and there is some variation among participating institutions in the definitions and diagnostic methods for PA subtypes (Supplementary Methods). Finally, our genomic mediation analysis assumes that the structural model is correctly specified and that there is no horizontal pleiotropy. Although we performed sensitivity analyses adjusting for metabolism-related covariates, the results may still be affected by model misspecification or unmeasured confounders.

In summary, this study advances our understanding of the genetic architecture of PA, provides genetic evidence that the elevated cardiovascular risk in PA is primarily driven by BP-independent mechanisms, and links polygenic aldosterone susceptibility to a continuous biological spectrum extending into the general population. These findings provide clues to the polygenic stratification of aldosterone-related diseases and highlight the potential of aldosterone-targeted therapies.

## Methods

### Japanese PA GWAS

We recruited patients with PA from three hospitals and a biobank network in Japan, collectively comprising the JPS cohort. PA was diagnosed according to the Japan Endocrine Society guidelines^43^, and subtypes were mainly classified using adrenal venous sampling (performed in 89.6% of cases) or a clinical prediction score^44^. Controls were drawn from BBJ^8,9^ and restricted to non-hypertensive individuals to minimize inclusion of undiagnosed PA. GWAS was performed using REGENIE^45^ with logistic regression and approximate Firth fallback, including age, sex, and the first ten principal components as covariates. Genome-wide significance was defined as *P* < 5.0 × 10^−8^. All participants provided written informed consent, and the study protocol was approved by the ethics committee of the Japan Institute for Health Security (JIHS) (application No. 004791). Details about case and control definition, phenotypic characterization, genotyping, imputation, and quality control procedures are described in the Supplementary Methods.

### Trans-Ancestry Meta-Analysis

External PA GWAS summary statistics were obtained from meta-analysis resources for large-scale cohorts (FinnGen^46^, Million Veteran Program (MVP)^47^ European, MVP African, and UK Biobank (UKBB)^48^, plus the precomputed European “leave_MVP_AFR” meta-analysis) and from the French COMETE cohort^5^ (GWAS Catalog GCST90129620). Datasets were converted to GRCh37 and restricted to autosomal biallelic SNPs with minor allele frequency (MAF) ≥ 0.01.

The primary trans-ancestry meta-analysis was performed using a fixed-effect IVW approach in METAL^49^ under a staged integration strategy: a European meta-analysis (COMETE + leave_MVP_AFR) was first assembled and then combined with the African (MVP African) and the Japanese (JPS) datasets to generate the final trans-ancestry results. As a secondary analysis, we performed trans-ethnic meta-regression with MR-MEGA^10^ on the six cohort-level datasets (FinnGen, MVP European, UKBB, COMETE, MVP African, and JPS). Additional information on the contributing studies and the MR-MEGA analysis is available in the Supplementary Methods.

Trans-ancestry lead SNPs used in subsequent analyses to represent each locus were based on the top-hit SNPs from the IVW meta-analysis, with the exception of *CLRN1* locus, where the MR-MEGA top-hit was selected. To assess cross-population concordance, we estimated the trans-ancestry genetic correlation between the European and the Japanese datasets using Popcorn^50^. In addition, we assessed heterogeneity in effect sizes for the trans-ancestry lead SNPs using Cochran’s *Q* and MR-MEGA, applying a Bonferroni-corrected threshold of *P* = 0.05/19 (the number of significant loci) = 2.63 × 10^−3^.

### Fine-Mapping and Functional Characterization

For each PA locus, we conducted trans-ancestry fine-mapping with SuSiEx^51^ in a ±500-kb window around the trans-ancestry lead SNP, using the 1000 Genomes Project (1kGP) reference panel. Variants within 95% credible sets were annotated using RegulomeDB^11,52^ and FORGEdb^12^.

At loci where the trans-ancestry lead SNP (or a linkage disequilibrium [LD] proxy with *r*^2^ ≥ 0.8 if unavailable) showed significant adrenal eQTL/sQTL association in the GTEx^13^ dataset (FDR ≤ 0.05), we conducted colocalization analyses between European PA and adrenal eQTL/sQTL signals using coloc^53^. A posterior probability of a shared causal variant (PP.H4) > 0.8 was considered strong evidence of colocalization. Furthermore, chromatin interaction mapping was performed on the FUMA^54^ SNP2GENE module under default settings, using adrenal Hi-C data (GEO accession GSE87112).

### Subtype-Stratified PA GWAS

In the JPS cohort, we performed subtype-stratified GWAS for APA and BAH, using the same controls and settings as in the primary PA GWAS. The results were then combined with the corresponding subtype-stratified datasets from the COMETE cohort (GWAS catalog: GCST90129623 and GCST90129624) using an IVW meta-analysis with METAL.

### Genetic Mediation Analysis

We conducted mediation analyses in Genomic SEM^55^ using the European PA dataset together with public GWAS summary statistics for SBP and four cardiovascular outcomes: CAD, AF, HF, and IS (Supplementary Table 1). We applied multivariable LDSC to derive the genetic covariance matrix (S) and corresponding sampling covariance matrix (V). We specified a structural mediation model that partitioned the total genetic effect of PA on each outcome into an SBP-mediated path (*a* and *b*) and an SBP-independent path (*c’*), and calculated the SBP-independent proportion as *c’* / (*ab* + *c’*). Identical mediation models were applied to the JPS datasets to evaluate inter-ancestry replication. Model sensitivity was evaluated in the European datasets by adding body mass index, glycated hemoglobin, triglycerides, and low-density lipoprotein cholesterol as covariates to the model (Supplementary Table 1; Supplementary Fig. 14).

### Inter-Trait Analysis between PA, SBP, and serum potassium

Based on previous findings indicating the presence of overlap between SBP and PA loci^4^, we utilized a list of loci strongly associated with SBP, derived from a previous BP GWAS conducted in East Asians^15^, to evaluate whether SBP-increasing alleles are also associated with increased risk of disease in this PA GWAS of Japanese individuals. As a reverse approach, we used public GWAS summary statistics to examine the associations of the 18 PA lead variants in Europeans (excluding *CLRN1*, which showed no association in Europeans) with SBP and serum potassium (Supplementary Table 1, Supplementary Methods: *Selection of European SBP Datasets*). Genetic correlations (*r*_g_) between PA and other traits were estimated with LDSC^14^ in both ancestries using European summary statistics. Additionally, we applied the LCV model^18^ to the European summary statistics to assess the causal relationships between the following pairs: PA and SBP, and PA and serum potassium. The direction of the causal effect estimate was set so that positive values correspond to the causal effect of PA on SBP or serum potassium.

### PA GRS Analysis

We constructed two ancestry-specific GRS models, tailored for the Japanese and European populations. For each model, we selected a subset of the trans-ancestry lead SNPs that were nominally significant (*P* < 0.05) within their respective ancestries, weighting them by their ancestry-specific effect sizes. From the Japanese model, we further derived a modified GRS that excluded the *TARID/TCF21* locus (rs36125724), which showed a distinct effect-size pattern between APA and BAH subtypes.

The standard Japanese GRS was applied to Japanese population reference cohorts (KING^19^ and BBJ cohorts), the European GRS to a subset of Europeans within the AoU Research Program^20^ data, and the modified Japanese GRS to Japanese PA cases (JPS). To enable comparisons between cohorts, individual scores for Japanese and European participants were standardized using the means and standard deviations of the KING and AoU cohorts, respectively, as reference values.

In population-based cohorts (KING, BBJ, and AoU), associations between the GRS and clinical traits, including SBP, serum potassium, and log-transformed PAC, were evaluated using univariate linear regression, excluding individuals receiving medications that influence these traits. In Japanese PA cases, associations with biochemical markers of aldosterone excess were evaluated using the modified GRS. Details about the construction and analyses of GRS are provided in the Supplementary Methods.

## Supporting information

Description of Additional Supplementary File

## Acknowledgements

We thank all study participants, investigators, and coordinators from the contributing cohorts. The sample and data used for this research were provided from the BioBank Japan Project that was supported by AMED. We gratefully acknowledge All of Us participants for their contributions, without whom this research would not have been possible. We also thank the National Institutes of Health’s All of Us Research Program for making available the participant data examined in this study. We also acknowledge the participants and investigators of the FinnGen, UK Biobank, Million Veteran Program, and KING studies.

## Author Contributions

Participant recruitment, characterization, and data generation. Yokohama Rosai Hospital cohort: Y.Tsurutani, T.N.; Tohoku University cohort: Y.Tezuka, Y.Y., Y.O., T.S., H.S., F.S.; Institute of Science Tokyo cohort: M.M., R.I., T.Y.; KING study cohort: M.N., M.Y., K.Y., S.I.; NCBN cohort: D.T., A.T., Y.H., N.K.; JPAS-II study registry: M.S. Statistical analyses: D.T., M.N., F.T., (with group lead, F. T.). Manuscript writing: N.K., D.T., F.T., M.N., M.S., A.T., F.S. (with group lead, N.K.). Study design and supervision: N.K. All authors critically reviewed and approved the final version of the manuscript.

## Data Availability

Summary statistics for the Japanese GWAS and trans-ancestry meta-analyses have been deposited in the GWAS Catalog under accession IDs GCST90985499–GCST90985504 [https://www.ebi.ac.uk/gwas/]. Individual-level genotype data are not publicly available because of privacy restrictions. Other data generated or analyzed in this study are available from the corresponding author upon reasonable request.

## Competing Interests

None

## Sources of funding

This study was supported by a grant (23A2010) from NCGM. The KING study was supported by JSPS KAKENHI Grant Numbers JP21H03206 and JP24K02690.

## Notes

### Competing Interest Statement

The authors have declared no competing interest.

### Author Declarations

Ethics committee of the Japan Institute for Health Security (JIHS) gave ethical approval for this work

