## Supplementary material for "Genetic Analysis of Primary Aldosteronism Bridging Disease Subtypes and the Phenotypic Continuum": Description of Additional Supplementary File

**Supplementary Data 1**
Description: Ancestry-specific associations and heterogeneity for lead SNPs at PA risk loci.

**Supplementary Data 2**
Description: Trans-ancestry fine-mapping and functional annotation of PA risk loci.

**Supplementary Data 3**
Description: Significant adrenal gland cis-eQTLs for lead SNPs at PA loci.

**Supplementary Data 4**
Description: Significant adrenal gland sQTLs for lead SNPs at PA risk loci.

**Supplementary Data 5**
Description: PA association statistics at the 56 top SBP-associated loci in East Asians.
