## Supplementary Information for "Genetic Analysis of Primary Aldosteronism Bridging Disease Subtypes and the Phenotypic Continuum"

**Tomidokoro *et al*.**

### Supplementary Figures

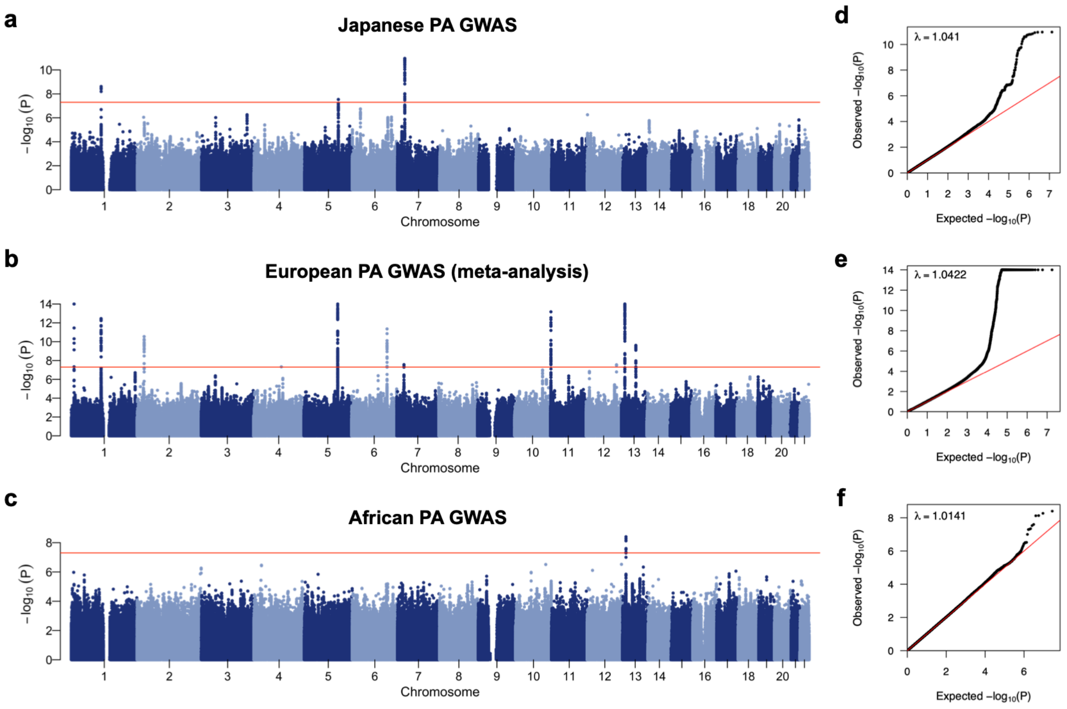

**Supplementary Figure 1. Genome-wide association study summary plots for primary aldosteronism across Japanese, European, and African populations. (a), (b), (c)** Manhattan plots for the Japanese **(a)**, European meta-analysis **(b)**, and African **(c)** primary aldosteronism (PA) genome-wide association study (GWAS) datasets. The y-axis denotes the negative $\log_{10}$ *P*-value, and the x-axis represents the chromosomal position. The horizontal red line marks the genome-wide significance threshold ($P < 5.0 \times 10^{-8}$). **(d), (e), (f)** Quantile-quantile (QQ) plots of the corresponding association results for Japanese **(d)**, European **(e)**, and African **(f)** datasets.

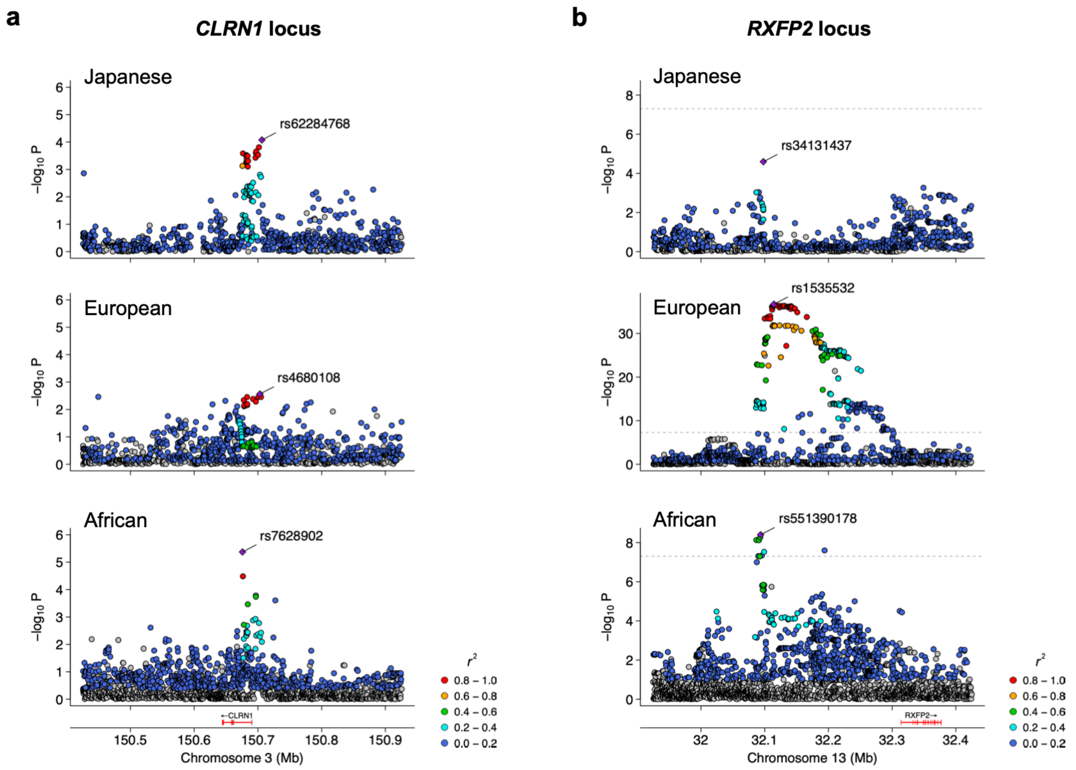

**Supplementary Figure 2. Ancestry-specific regional association patterns at the *CLRN1* and *RXFP2* loci. (a)** Regional plots at the *CLRN1* locus showing primary aldosteronism (PA) association signals in Japanese (top), European (middle), and African (bottom) populations, and **(b)** regional plots at the *RXFP2* locus comparing signals across the same set of ancestries. Points are colored by linkage disequilibrium ($r^2$) with population-specific lead variants.

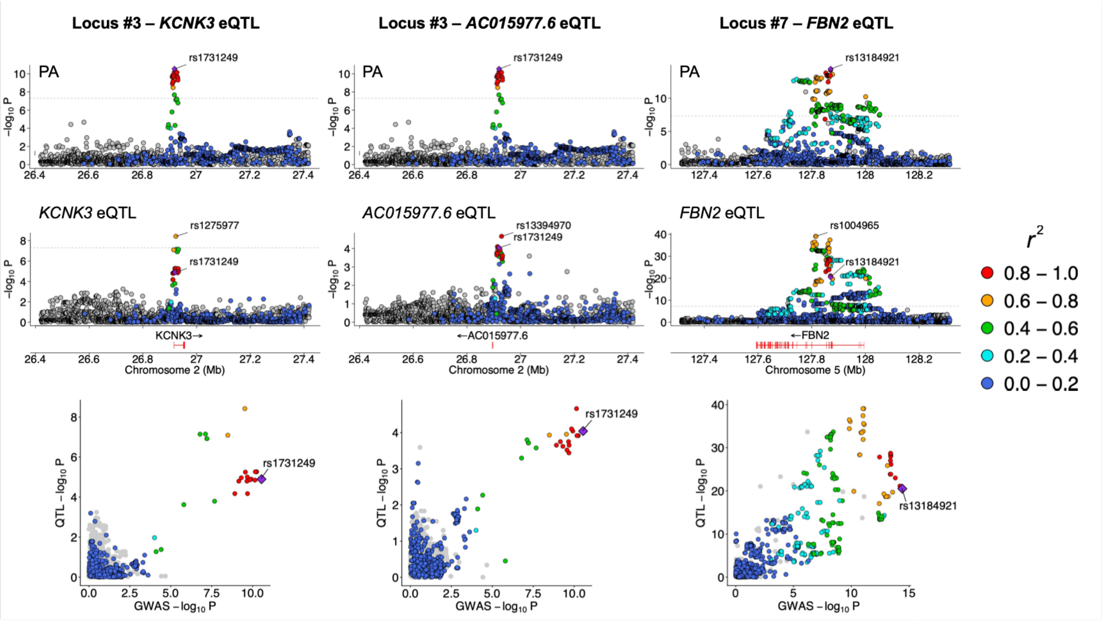
Locus #3 – KCNK3 eQTL
Locus #3 – AC015977.6 eQTL
Locus #7 – FBN2 eQTL
PA
rs1731249
rs13184921
KCNK3 eQTL
AC015977.6 eQTL
FBN2 eQTL
rs1275977
rs13394970
rs1004965
KCNK3→
←AC015977.6
←FBN2
Chromosome 2 (Mb)
Chromosome 5 (Mb)
QTL –log10 P
GWAS –log10 P
r2
0.8 – 1.0
0.6 – 0.8
0.4 – 0.6
0.2 – 0.4
0.0 – 0.2

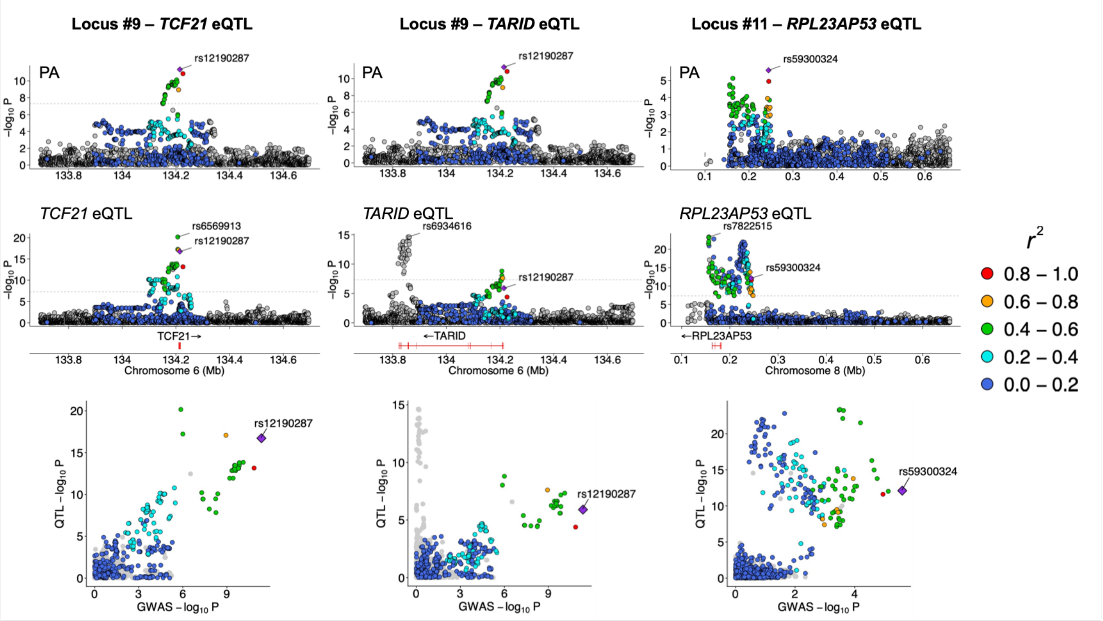
Locus #9 – TCF21 eQTL
Locus #9 – TARID eQTL
Locus #11 – RPL23AP53 eQTL
PA
rs12190287
rs59300324
TCF21 eQTL
TARID eQTL
RPL23AP53 eQTL
rs6569913
rs6934616
rs7822515
TCF21→
←TARID
←RPL23AP53
Chromosome 6 (Mb)
Chromosome 8 (Mb)
QTL –log10 P
GWAS –log10 P
r2
0.8 – 1.0
0.6 – 0.8
0.4 – 0.6
0.2 – 0.4
0.0 – 0.2

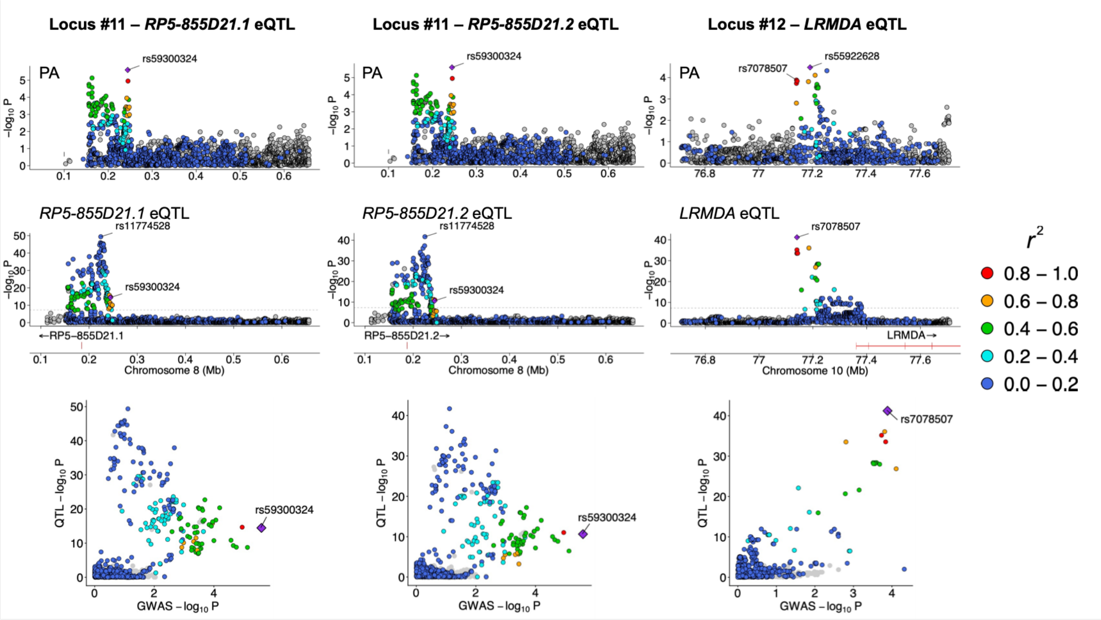

Locus #11 – RP5-855D21.1 eQTL
Locus #11 – RP5-855D21.2 eQTL
Locus #12 – LRMDA eQTL
PA
rs59300324
rs7078507
rs55922628
RP5-855D21.1 eQTL
RP5-855D21.2 eQTL
LRMDA eQTL
rs11774528
←RP5-855D21.1
RP5-855D21.2→
LRMDA→
Chromosome 8 (Mb)
Chromosome 10 (Mb)
QTL –log10 P
GWAS –log10 P
−log10 P
r2
0.8 – 1.0
0.6 – 0.8
0.4 – 0.6
0.2 – 0.4
0.0 – 0.2

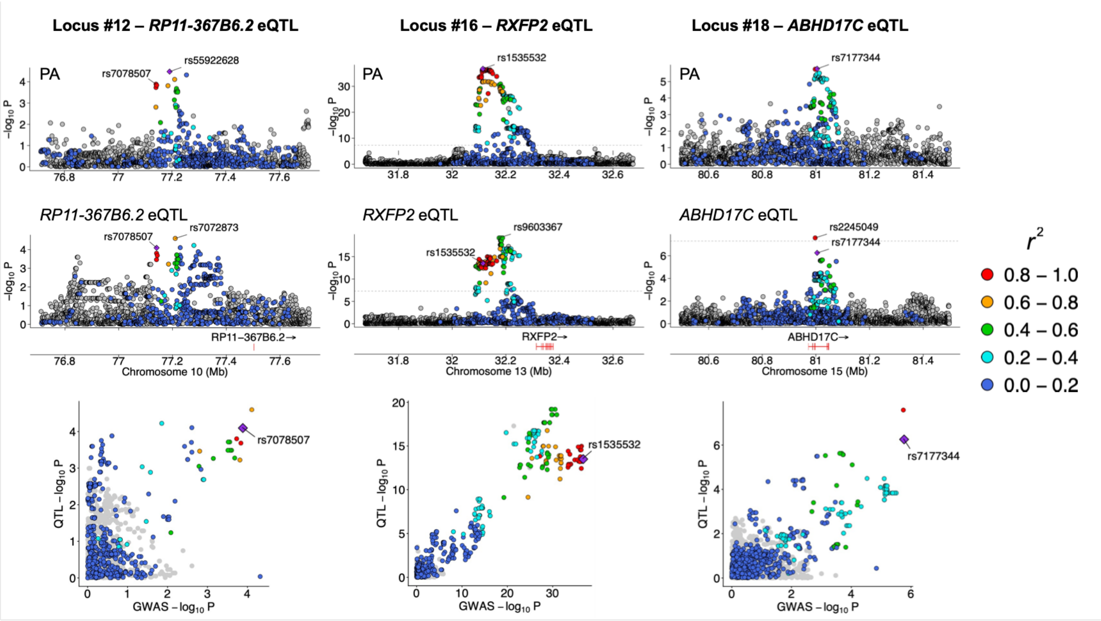

Locus #12 – RP11-367B6.2 eQTL
Locus #16 – RXFP2 eQTL
Locus #18 – ABHD17C eQTL
PA
rs7078507
rs55922628
rs1535532
rs7177344
RP11-367B6.2 eQTL
RXFP2 eQTL
ABHD17C eQTL
rs7072873
rs9603367
rs2245049
RP11−367B6.2→
RXFP2→
ABHD17C→
Chromosome 10 (Mb)
Chromosome 13 (Mb)
Chromosome 15 (Mb)
QTL –log10 P
GWAS –log10 P
−log10 P
r2
0.8 – 1.0
0.6 – 0.8
0.4 – 0.6
0.2 – 0.4
0.0 – 0.2

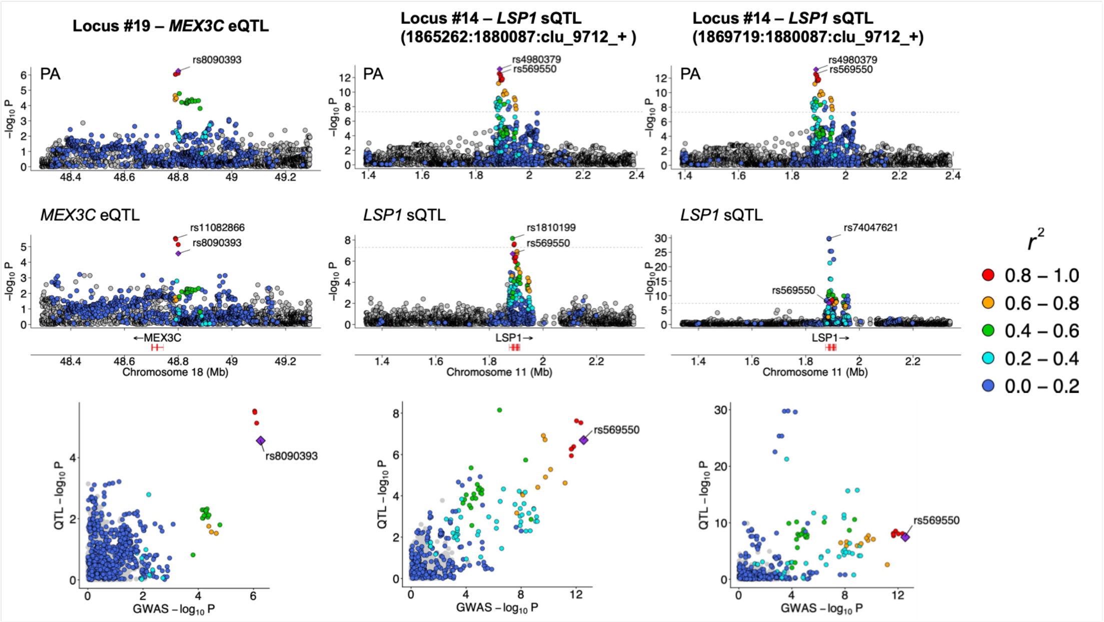

**Supplementary Figure 3. Colocalization of PA-associated loci with adrenal eQTLs or sQTLs.** For each locus, illustrations are presented in a three-tiered layout consisting of top, middle, and bottom panels. The top panels show regional plots of PA associations, with the European PA lead SNP annotated. The middle panels display the corresponding eQTL/sQTL associations, with both the eQTL/sQTL lead SNP and the PA lead SNP annotated in European populations. The bottom panels present scatter plots comparing association *P*-values between PA and eQTL/sQTL signals. Across all panels, data points are colored according to linkage disequilibrium ($r^2$) with the European PA lead SNP. For loci #12 and #14, where the European PA lead SNPs (rs55922628 and rs4980379, respectively) are absent from the eQTL/sQTL dataset, proxy SNPs were used. In these cases, the top panels are annotated with the PA lead SNP and its proxy, and the middle panels are annotated with the eQTL/sQTL lead SNP and the proxy. Color coding in the middle and bottom panels is defined relative to the proxy SNP for these loci. QTL, quantitative trait locus.

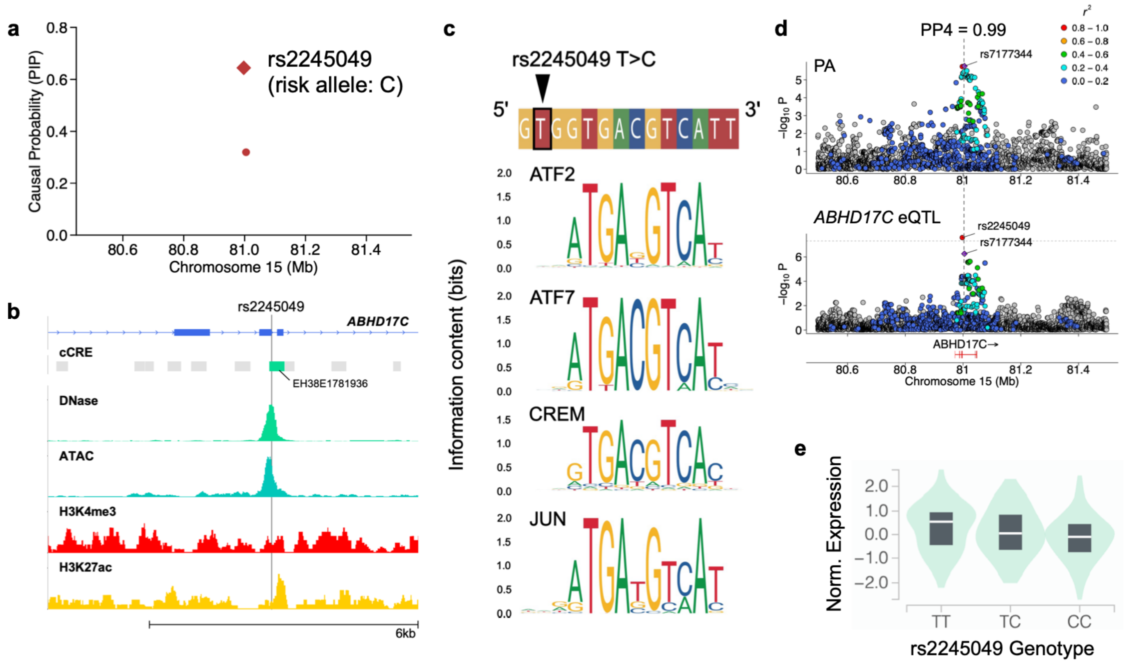

**Supplementary Figure 4. Functional prioritization of rs2245049 at the *ABHD17C* locus.** **(a)** Regional fine-mapping plot displaying the posterior inclusion probability (PIP) for each variant in the credible set identified at the *ABHD17C* locus. The top SuSiEx SNP, rs2245049 (PIP = 0.64), shows the strongest regulatory evidence across annotation databases, with a RegulomeDB score of 1a and a FORGEdb score of 10. **(b)** Functional genomic annotations at the *ABHD17C* locus. The rs2245049 variant overlaps an active cis-regulatory element (cCRE) defined by open chromatin (DNase, ATAC) and active histone marks (H3K4me3, H3K27ac). **(c)** Sequence motif analysis showed that rs2245049 overlaps the 5' regulatory footprint of transcription factors ATF2, ATF7, CREM, and JUN, suggesting a potential role in modulating their binding affinity. ATF2/7 and CREB1 are known to mediate signaling pathways that regulate *CYP11B2* expression in response to angiotensin II and potassium. **(d)** Regional plots comparing the association signals for primary aldosteronism (PA; top) and *ABHD17C* mRNA expression (bottom) in the European population. Data points are colored according to $r^2$ with the European PA lead SNP rs7177344. Colocalization analysis showed that the genetic signal for PA overlaps with an eQTL for *ABHD17C* in the adrenal gland (PP.H4 = 0.99), suggesting a shared underlying variant. **(e)** Violin plots illustrating normalized *ABHD17C* expression stratified by rs2245049 genotype, indicating that the PA risk-associated C allele correlates with a dose-dependent decrease in *ABHD17C* mRNA expression.

**a**

**b**

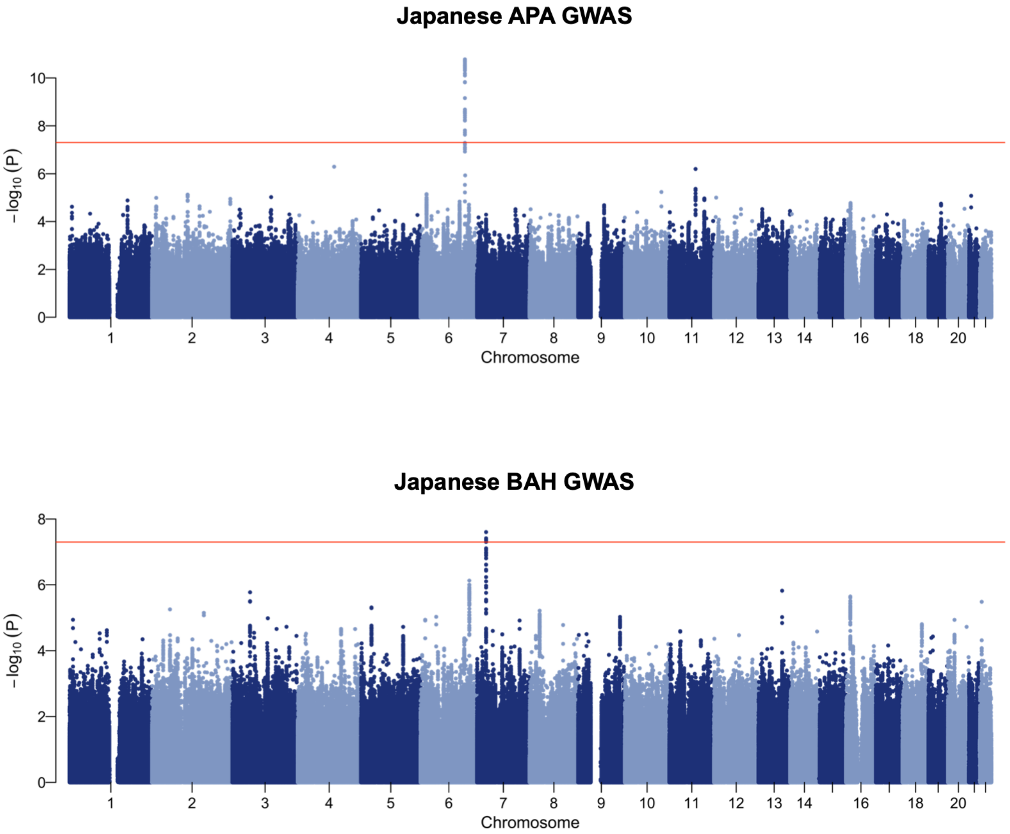

**Supplementary Figure 5. GWAS for PA subtypes in the Japanese cohort.** Manhattan plots displaying the genome-wide association results for two subtypes of primary aldosteronism (PA) in the Japanese cohort. **(a)** Results for aldosterone-producing adenoma (APA). **(b)** Results for bilateral adrenal hyperplasia (BAH). The red horizontal lines indicate the genome-wide significance level ($P = 5.0 \times 10^{-8}$).

a

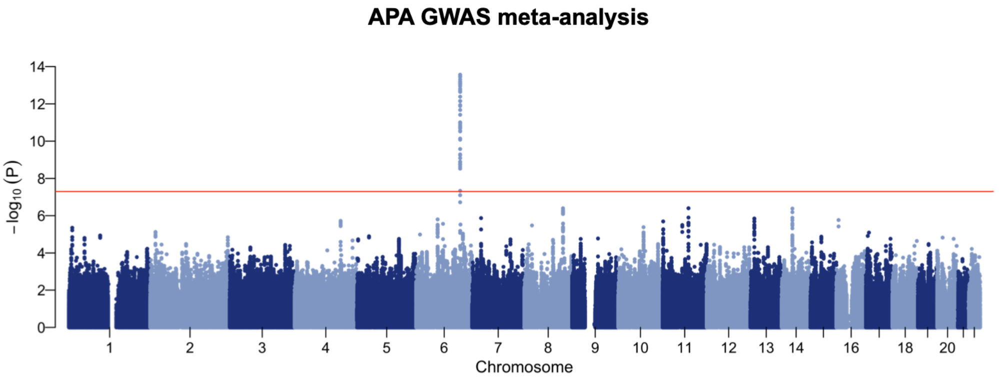

b

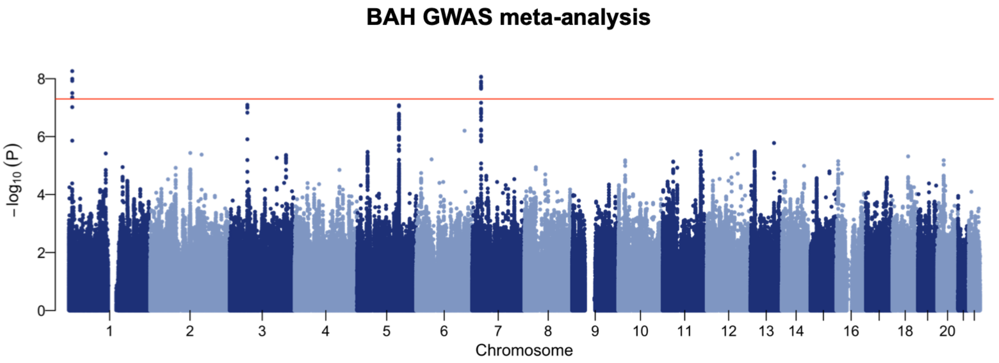

**Supplementary Figure 6. Meta-analysis of subtype-stratified PA GWAS.** Manhattan plots for the combined subtype-stratified PA GWAS meta-analyses combining the Japanese JPS and French COMETE cohorts. **(a)** Results for aldosterone-producing adenoma (APA). **(b)** Results for bilateral adrenal hyperplasia (BAH). The red horizontal lines indicate the genome-wide significance level ($P = 5.0 \times 10^{-8}$).

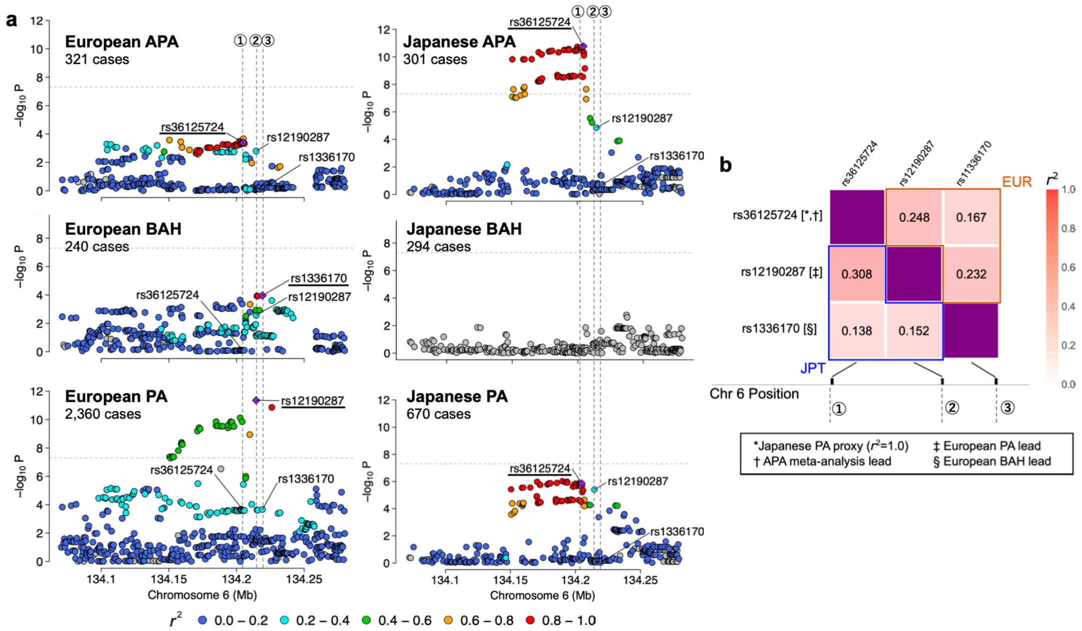

**Supplementary Figure 7. Distinct ancestry- and subtype-specific genetic architectures identified at the *TARID/TCF21* locus. (a)** Regional plots at the *TARID/TCF21* locus across European and Japanese cohorts for APA (top), BAH (middle), and overall PA (bottom). Point colors indicate $r^2$ with the index SNP for each group, denoted by a purple diamond and an underlined SNP label. The association signals across these groups are captured by three index SNPs: rs36125724 (①), rs12190287 (②), and rs1336170 (③). In Europeans, the associations separate into distinct signals: rs1336170 (③) is the BAH lead, rs12190287 (②) is the PA lead, and rs36125724 (①) serves as a proxy for the APA signal. Conversely, the association results in the Japanese cohort lack a significant BAH signal; the PA association in Japanese cohort is primarily driven by the APA lead SNP rs36125724 (①). **(b)** Linkage disequilibrium (LD) matrices displaying $r^2$ values among the three representative SNPs in European (EUR; the upper-right part) and Japanese (JPT; the lower-left part) populations. In Europeans, the European PA lead SNP rs12190287 shows similar LD with both the APA meta-analysis lead (rs36125724; $r^2$ = 0.25) and European BAH lead (rs1336170; $r^2$ = 0.23) SNPs. The latter two SNPs are themselves in weak LD ($r^2$ = 0.17), supporting the presence of independent subtype-specific signals in this population.

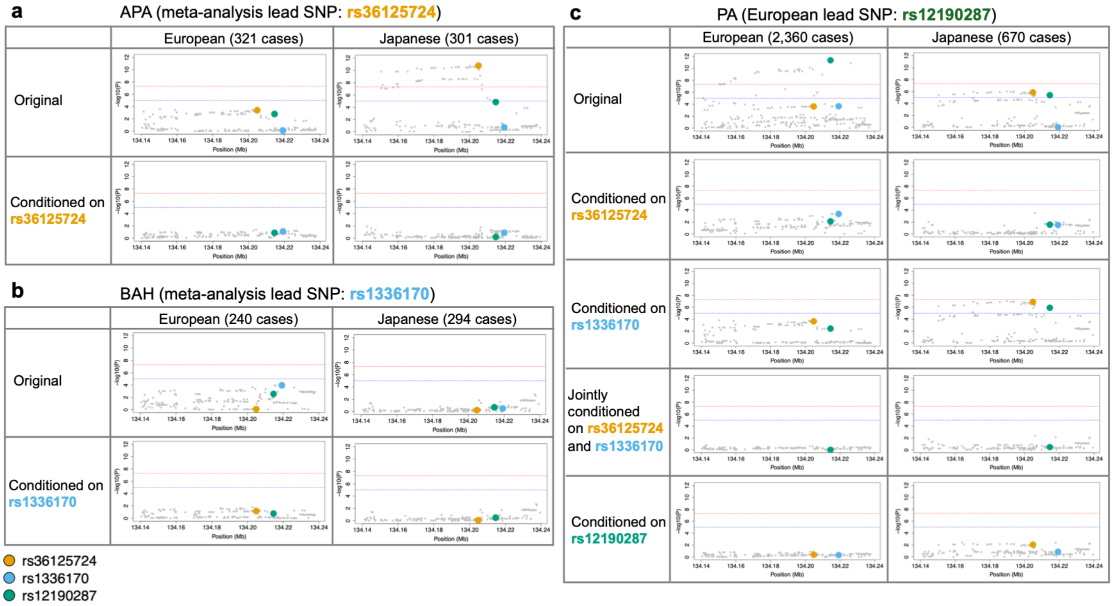

**Supplementary Figure 8. Fine-mapping of association signals at the *TARID/TCF21* locus with PA as a whole and its subtypes. (a)** Conditional analysis of the aldosterone-producing adenoma (APA) signal in European (left) and Japanese (right) cohorts. The top row shows unconditioned results, with the meta-analysis lead SNP, rs36125724, indicated by an orange point. Conditioning on this SNP completely eliminates the signal in both populations. **(b)** Conditional analysis of the bilateral adrenal hyperplasia (BAH) signal in the same cohorts. The top row shows unconditioned results, with the lead SNP, rs1336170, indicated by a blue point. Conditioning on rs1336170 leads to a noticeable reduction in the signal in Europeans, but in the Japanese cohort, where no primary BAH signal is observed at baseline, the change is minimal. **(c)** Conditional analysis of overall PA. The top row shows baseline unconditioned analysis; subsequent rows show analyses conditioned on the APA lead SNP (rs36125724, orange point), the BAH lead SNP (rs1336170, blue point), both variants simultaneously, and the European PA lead SNP (rs12190287, green point) (refer to Supplementary Fig. 7). In Europeans, the PA signal is partially attenuated when conditioned on only one of the SNPs, rs36125724 or rs1336170, but it disappears completely when conditioned on both SNPs. This suggests that the PA signal in Europeans reflects a composite of subtype-specific associations. Furthermore, the PA signal is also weakened when conditioned solely on the primary SNP, rs12190287. In contrast, in Japanese individuals, conditioning on the APA lead SNP rs36125724 alone largely eliminates the PA signal, reflecting the dominance of the APA component in this population.

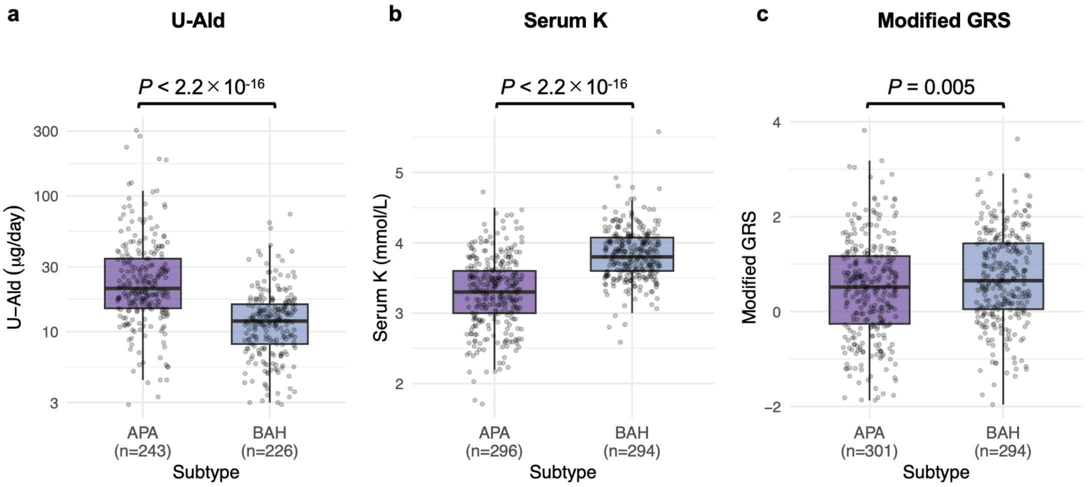

**Supplementary Figure 9. Comparison of biochemical measurements and modified genetic risk scores between PA subtypes.** Boxplots comparing aldosterone-producing adenoma (APA) and bilateral adrenal hyperplasia (BAH) for three variables: **(a)** 24-hour urinary aldosterone excretion (U-Ald, µg/day, log-scale), **(b)** serum potassium level (mmol/L), and **(c)** modified PA GRS. The modified PA GRS excludes the *TARID/TCF21* locus to account for its disproportionate effect on APA susceptibility.

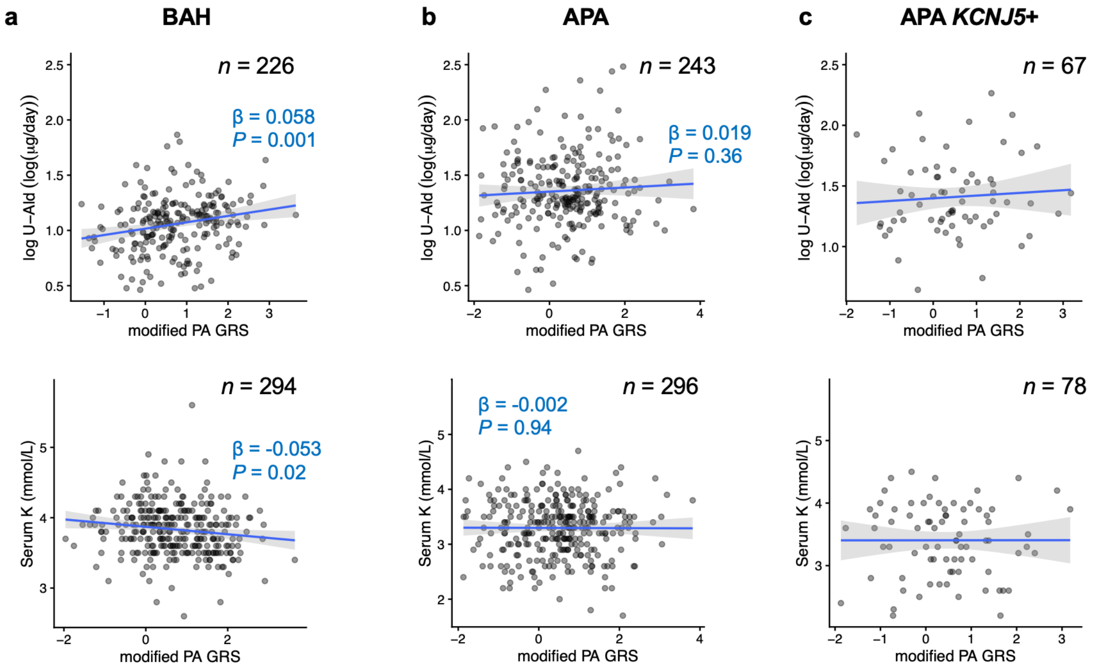

**Supplementary Figure 10. Correlation between modified PA GRS and biochemical measurements across PA subtypes.** Scatter plots displaying the modified GRS plotted against log-transformed 24-hour urinary aldosterone excretion (top row) and serum potassium level (bottom row). The data are stratified into three subgroups: **(a)** bilateral adrenal hyperplasia (BAH), **(b)** aldosterone-producing adenoma (APA), and **(c)** APA with a *KCNJ5* somatic mutation. Solid lines show the approximation obtained by linear regression, and the shaded areas represent the 95% confidence interval. Beta coefficients and *P*-values are derived from unadjusted linear regression models.

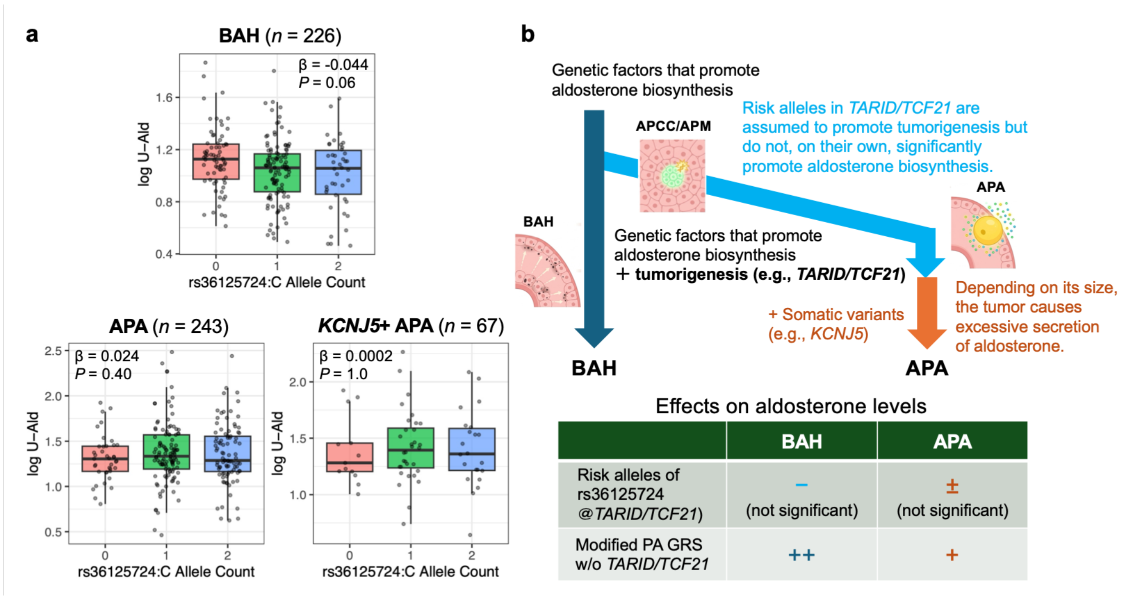

**Supplementary Figure 11. Genetic influence of *TARID/TCF21* rs36125724: correlation between genotype and 24-hour urinary aldosterone excretion, and the "two-step hit" hypothesis. (a)** The graphs show the relationship between the number of risk alleles for rs36125724 and 24-hour urinary aldosterone excretion (U-Ald) in the BAH group (*n* = 226, top), the APA group (*n* = 243, lower left), and the subgroup of APA carrying a KCNJ5 somatic mutation (*n* = 67, lower right). Beta coefficients and *P*-values are derived from multivariable linear regression models adjusted for age, sex, and mGRS. No significant positive correlation was observed in any of these groups. **(b)** A schematic diagram of the "two-step hit" hypothesis that can explain the findings regarding *TCF21* identified in this study. As the disease progresses from BAH to APCC/APM and then to APA, genetic factors involved in aldosterone biosynthesis other than the *TARID*/*TCF21* locus may influence all of these conditions. On the other hand, while risk alleles at the *TARID*/*TCF21* ("first-hit") locus increase tumorigenicity, the *TARID*/*TCF21* locus alone does not appear to have a significant effect on aldosterone biosynthesis. It is hypothesized that when somatic mutations ("second hits") occur in genes that cause APA, such as *KCNJ5*, in addition to *TCF21*, tumorigenicity is further promoted, leading to the production of excessive aldosterone as APA. However, in APA, the amount of aldosterone synthesized is greatly influenced by tumor size; therefore, the impact of other genetic factors in APA (such as the modified GRS excluding *TCF21*) is smaller than that in BAH (bottom table).

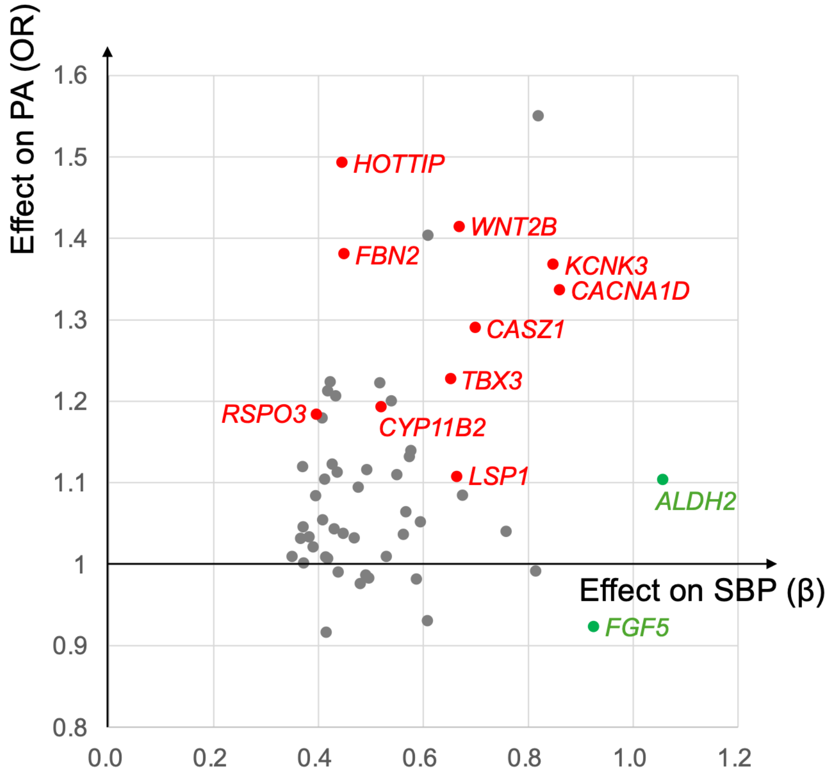

**Supplementary Figure 12. Genetic overlap of SBP and PA in East Asians.** The scatter plot compares the effect sizes on SBP and the odds ratios for PA at the top 56 SBP-associated loci in East Asians[1]. The red points indicate SBP loci that are also significantly associated with PA in our trans-ancestry PA GWAS meta-analysis. These loci are shown alongside *CYP11B2*, which is well-known for its biological significance in PA. The green points indicate *ALDH2* and *FGF5*, which have a large effect on SBP but show only a weak association with PA. This divergence is consistent with previous studies implicating these loci in blood pressure pathways that appear to be independent of adrenal or aldosterone activity[2,3].

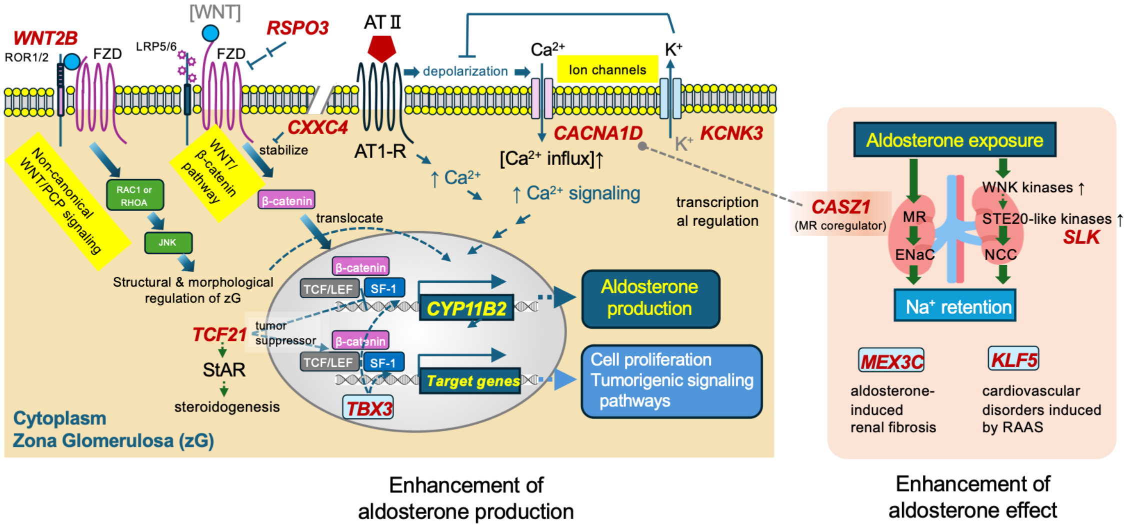

**Supplementary Figure 13. Biological pathways involving target genes inferred from fine mapping of the PA loci.** Many of the identified target genes (see Table 1) are components of pathways involved in aldosterone biosynthesis and function.

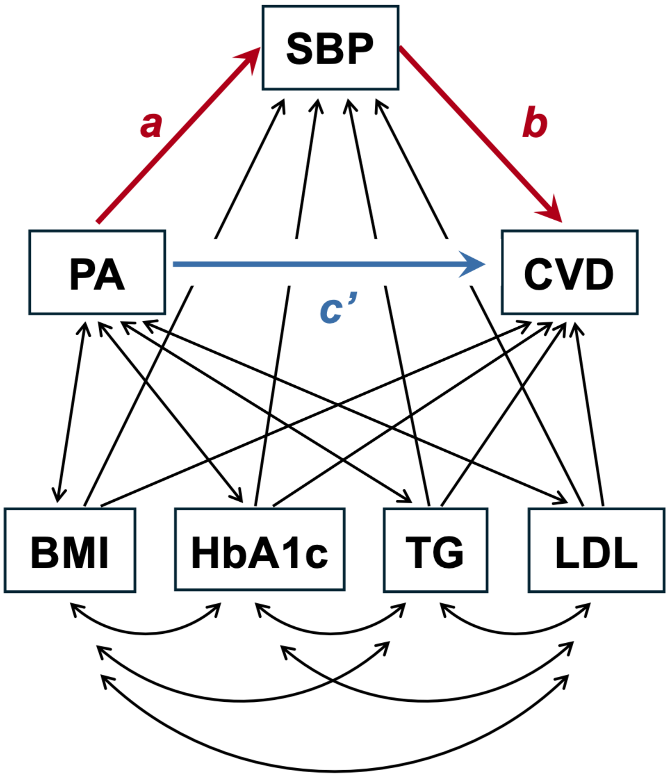

**Supplementary Figure 14. Extended structural mediation model incorporating metabolic covariates.** Schematic diagram of the model used in sensitivity testing of the primary mediation analysis. The structural model specifies directional paths from PA and metabolic covariates (body mass index [BMI], glycated hemoglobin [HbA1c], triglycerides [TG], and low-density lipoprotein [LDL]) to systolic blood pressure (SBP). Similarly, directional paths are specified from PA, SBP, and the metabolic covariates to cardiovascular disease (CVD). PA and the metabolic covariates are allowed to covary, as are the covariates with one another.

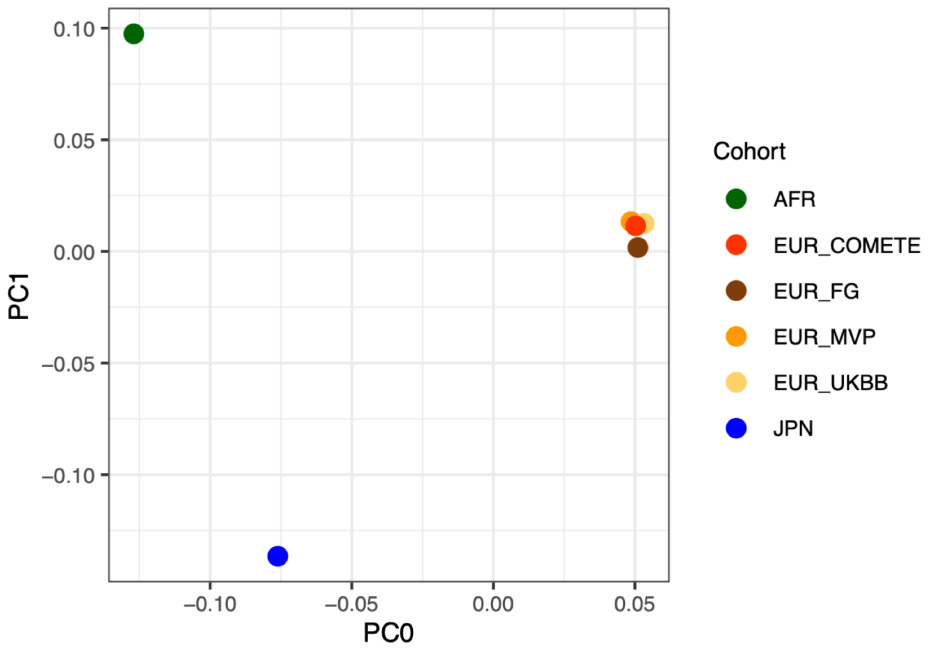

**Supplementary Figure 15. Axes of genetic variation across the study cohorts.** The scatter plot shows the first two principal components (PC0 and PC1) generated by MR-MEGA, derived from multi-dimensional scaling of allele frequency differences. Colors indicate the respective demographic cohorts across the African (AFR), European (EUR: COMETE, FinnGen [FG], MVP, and UKBB), and Japanese (JPN) datasets.

### Supplementary Tables

#### Supplementary Table 1. Data sources for publicly available GWAS summary statistics.

| Ancestry | Trait | Data Source (Identifier) | PMID | Analysis Performed |
|---|---|---|---|---|
| European | SBP (adjusted for BMI) | GWAS Catalog (GCST90310294) | 38689001 | Locus-level comparison |
| | SBP (non-adjusted for BMI) | GWAS Catalog (GCST90691848) | 40968291 | Genetic mediation<br>Genetic correlation<br>LCV model |
| | Serum potassium | GWAS Catalog (GCST90476307) | 39024449 | Locus-level comparison<br>Genetic correlation<br>LCV model |
| | Coronary artery disease | GWAS Catalog (GCST90132314) | 36474045 | Genetic mediation |
| | Atrial fibrillation | GWAS Catalog (GCST006414) | 30061737 | Genetic mediation |
| | Heart failure | GWAS Catalog (GCST90728695) | 40038546 | Genetic mediation |
| | Ischemic stroke | GWAS Catalog (GCST90104540) | 36180795 | Genetic mediation |
| | Body mass index | GWAS Catalog (GCST90029007) | 29892013 | Genetic mediation<br>(Covariate) |
| | HbA1c | GWAS Catalog (GCST90691576) | 40968291 | Genetic mediation<br>(Covariate) |
| | Triglycerides | GWAS Catalog (GCST90239664) | 34887591 | Genetic mediation<br>(Covariate) |
| | Low-density lipoprotein | GWAS Catalog (GCST90239658) | 34887591 | Genetic mediation<br>(Covariate) |
| Japanese<br>/East Asian | SBP | GWAS Catalog (GCST90278668) | 38116116 | Genetic mediation<br>Genetic correlation<br>LCV model |
| | Serum potassium | Jenger database (QTL GWAS ID 229) | 39363016 | Genetic correlation<br>LCV model |
| | Coronary artery disease | NBDC database (hum0014.v20.cad.v1) | 33020668 | Genetic mediation |
| | Atrial fibrillation | NBDC database (hum0014.v29.AF.v1) | 36653681 | Genetic mediation |
| | Heart failure | GWAS Catalog (GCST90668009) | 41184235 | Genetic mediation |
| | Ischemic stroke | GWAS Catalog (GCST90018644) | 34594039 | Genetic mediation |

**Supplementary Table 2. Demographics and clinical characteristics of the case and control cohorts in the Japanese GWAS.**

| | PA cases (JPS cohort) | | | | Non-hypertensive BBJ Control (*n* = 25,998) |
|---|---|---|---|---|---|
| | Total (*n* = 670) | Subtypes | | | |
| | | APA (*n* = 301) | BAH (*n* = 294) | Inconclusive (*n* = 75) | |
| **Demographics** | | | | | |
| Age (years), mean (SD) | 52.6 (11.3) | 52.3 (11.3) | 52.4 (11.5) | 54.2 (10.3) | 62.4 (14.3) |
| Male sex, *n* (%) | 325 (48.5) | 159 (52.8) | 122 (41.5) | 44 (58.7) | 12,081 (46.5) |
| **Clinical parameters** | | | | | |
| PRA (ng/mL/h), median [IQR] | 0.30 [0.20, 0.45] | 0.20 [0.20, 0.40] | 0.30 [0.20, 0.50] | 0.28 [0.20, 0.40] | |
| PAC (ng/dL), median [IQR] | 22.5 [15.3, 35.5] | 32.6 [23.0, 52.7] | 17.1 [13.7, 24.0] | 17.1 [12.7, 26.1] | |
| ARR (ng/dL per ng/mL/h), median [IQR] | 80.5 [45.0, 155.4] | 148.8 [72.9, 297.4] | 57.2 [33.6, 89.6] | 76.1 [38.8, 103.5] | N/A |
| U-Ald (μg/24h), median [IQR] | 15.0 [10.1, 23.6] | 20.8 [14.9, 34.5] | 12.0 [8.1, 15.9] | 9.3 [6.2, 15.0] | |
| Serum potassium (mmol/L), mean (SD) | 3.55 (0.51) | 3.30 (0.51) | 3.83 (0.36) | 3.47 (0.49) | |

**Supplementary Table 3. Genome-wide significant loci associated with PA in the Japanese cohort.**

| Prioritized Gene | Lead SNP | Chr | Position (GRCh37) | RA/NRA | Case RAF | Control RAF | OR (95% CI) | *P*-value |
|---|---|---|---|---|---|---|---|---|
| *WNT2B* | rs12038119 | 1 | 113043423 | G/A | 0.37 | 0.29 | 1.43 (1.27–1.60) | $2.4 \times 10^{-9}$ |
| *FBN2* | rs6595829 | 5 | 127817651 | A/C | 0.32 | 0.24 | 1.42 (1.26–1.59) | $2.9 \times 10^{-8}$ |
| *HOTTIP* | rs3735533 | 7 | 27245893 | C/T | 0.68 | 0.59 | 1.50 (1.33–1.68) | $1.1 \times 10^{-11}$ |

**Supplementary Table 4. Study populations and sample sizes for the trans-ancestry meta-analysis.**

| Cohort | Ancestry | Cases (*n*) | | | Controls (*n*) |
|---|---|---|---|---|---|
| | | Total PA | APA | BAH | |
| JPS | Japanese | 670 | 301 | 294 | 25,998 |
| FinnGen | European | 931 | – | | 479,069 |
| UKBB | | 147 | – | | 413,289 |
| MVP European | | 720 | – | | 457,510 |
| COMETE | | 562 | 321 | 240 | 950 |
| MVP African | African | 676 | – | | 120,849 |
| Total | | 3,706 | 622 | 534 | 1,497,665 |

**Supplementary Table 5. Genome-wide significant loci identified through inverse-variance weighted trans-ancestry meta-analysis using METAL.**

| Locus ID | Prioritized Gene | Lead SNP | Chr | Position (GRCh37) | RA/NRA | OR (95% CI) | *P*-value |
|---|---|---|---|---|---|---|---|
| Locus #1 | *CASZ1* | rs880315 | 1 | 10796866 | C/T | 1.30 (1.24–1.37) | $1.8 \times 10^{-24}$ |
| Locus #2 | *WNT2B* | rs10776752 | 1 | 113044328 | T/G | 1.38 (1.29–1.48) | $2.2 \times 10^{-21}$ |
| Locus #3 | *KCNK3* | rs1731249 | 2 | 26920025 | T/A | 1.23 (1.17–1.29) | $5.1 \times 10^{-15}$ |
| Locus #4 | *CACNA1D* | rs3821843 | 3 | 53558012 | A/G | 1.19 (1.13–1.26) | $1.8 \times 10^{-10}$ |
| Locus #6 | *CXXC4* | rs71599015 | 4 | 105634811 | A/G | 1.66 (1.39–2.00) | $4.6 \times 10^{-8}$ |
| Locus #7 | *FBN2* | rs1004965 | 5 | 127814937 | C/T | 1.28 (1.22–1.35) | $2.3 \times 10^{-21}$ |
| Locus #8 | *RSPO3* | rs4897196 | 6 | 127225202 | T/A | 1.17 (1.11–1.23) | $2.3 \times 10^{-9}$ |
| Locus #9 | *TARID/TCF21* | rs12192720 | 6 | 134195719 | G/A | 1.26 (1.20–1.33) | $1.8 \times 10^{-17}$ |
| Locus #10 | *HOTTIP* | rs3735533 | 7 | 27245893 | C/T | 1.45 (1.34–1.57) | $4.0 \times 10^{-19}$ |
| Locus #11 | *RPL23AP53* | rs71512836 | 8 | 156284 | A/C | 1.20 (1.12–1.28) | $2.6 \times 10^{-8}$ |
| Locus #12 | *LRMDA* | rs7072873 | 10 | 77210191 | C/T | 1.15 (1.09–1.21) | $3.1 \times 10^{-8}$ |
| Locus #13 | *SLK* | rs9419958 | 10 | 105675946 | T/C | 1.24 (1.16–1.33) | $6.1 \times 10^{-10}$ |
| Locus #14 | *LSP1* | rs4980379 | 11 | 1888614 | T/C | 1.24 (1.17–1.30) | $6.9 \times 10^{-16}$ |
| Locus #15 | *TBX3* | rs35429 | 12 | 115555867 | A/G | 1.20 (1.14–1.26) | $6.0 \times 10^{-12}$ |
| Locus #16 | *RXFP2* | rs9603367 | 13 | 32175002 | T/C | 1.41 (1.33–1.49) | $5.0 \times 10^{-34}$ |
| Locus #17 | *KLF5* | rs78716205 | 13 | 73832307 | A/G | 1.54 (1.36–1.74) | $1.4 \times 10^{-11}$ |
| Locus #18 | *ABHD17C* | rs2245049 | 15 | 80997197 | C/T | 1.17 (1.11–1.24) | $6.8 \times 10^{-9}$ |
| Locus #19 | *MEX3C* | rs11082867 | 18 | 48790842 | C/A | 1.18 (1.12–1.25) | $1.0 \times 10^{-8}$ |

**Supplementary Table 6. Genome-wide significant loci identified through trans-ancestry meta-regression using MR-MEGA.**

| Locus ID | Prioritized Gene | Lead SNP | Chr | Position (GRCh37) | RA | A1/A2 | $\beta_0$ | $SE_0$ | $\beta_1$ | $SE_1$ | $\beta_2$ | $SE_2$ | *P*-value |
|---|---|---|---|---|---|---|---|---|---|---|---|---|---|
| Locus #1 | *CASZ1* | rs880315 | 1 | 10796866 | C | C/T | 0.26 | 0.05 | 0.38 | 0.65 | -0.13 | 0.68 | $9.2 \times 10^{-23}$ |
| Locus #2 | *WNT2B* | rs10776752 | 1 | 113044328 | T | T/G | 0.30 | 0.02 | 0.53 | 0.32 | -0.66 | 0.29 | $8.6 \times 10^{-20}$ |
| Locus #3 | *KCNK3* | rs1731249 | 2 | 26920025 | T | A/T | -0.21 | 0.01 | -0.11 | 0.13 | 0.86 | 0.15 | $3.6 \times 10^{-14}$ |
| Locus #4 | *CACNA1D* | rs3821843 | 3 | 53558012 | A | A/G | 0.17 | 0.05 | -0.50 | 0.69 | -0.41 | 0.65 | $1.5 \times 10^{-9}$ |
| Locus #5 | *CLRN1* | rs7628902 | 3 | 150675697 | T | C/T | -0.02 | 0.01 | 2.17 | 0.18 | 0.16 | 0.17 | $3.3 \times 10^{-8}$ |
| Locus #7 | *FBN2* | rs1004965 | 5 | 127814937 | C | C/T | 0.24 | 0.02 | -0.31 | 0.21 | -0.58 | 0.21 | $4.4 \times 10^{-20}$ |
| Locus #8 | *RSPO3* | rs12663110 | 6 | 127219931 | C | C/T | 0.15 | 0.03 | -0.73 | 0.41 | -0.07 | 0.43 | $1.1 \times 10^{-8}$ |
| Locus #9 | *TARID/TCF21* | rs12192720 | 6 | 134195719 | G | A/G | -0.23 | 0.03 | -0.02 | 0.42 | 0.03 | 0.40 | $1.3 \times 10^{-15}$ |
| Locus #10 | *HOTTIP* | rs3735533 | 7 | 27245893 | C | C/T | 0.35 | 0.04 | -0.25 | 0.70 | -0.25 | 0.59 | $2.3 \times 10^{-17}$ |
| Locus #13 | *SLK* | rs9419958 | 10 | 105675946 | T | C/T | 0.19 | 0.35 | -6.33 | 5.18 | -12.27 | 10.28 | $9.0 \times 10^{-9}$ |
| Locus #14 | *LSP1* | rs4980379 | 11 | 1888614 | T | T/C | 0.21 | 0.03 | 0.50 | 0.43 | 0.03 | 0.43 | $2.4 \times 10^{-14}$ |
| Locus #15 | *TBX3* | rs35436 | 12 | 115554523 | C | T/C | -0.18 | 0.02 | -0.04 | 0.25 | 0.22 | 0.27 | $2.5 \times 10^{-10}$ |
| Locus #16 | *RXFP2* | rs627879 | 13 | 32099925 | C | A/C | -0.32 | 0.05 | -1.45 | 0.60 | -1.47 | 0.60 | $9.2 \times 10^{-37}$ |
| Locus #17 | *KLF5* | rs78716205 | 13 | 73832307 | A | A/G | 0.30 | 0.43 | 1.72 | 6.52 | 4.55 | 12.63 | $5.3 \times 10^{-10}$ |

**Supplementary Table 7. Notable regulatory evidence for credible set variants in the adrenal gland.**

| Locus ID | Prioritized Gene | SNP | SuSiEx PIP | RegulomeDB | | FORGEdb Score | Summary of Regulatory Evidence |
|---|---|---|---|---|---|---|---|
| | | | | Rank | Adrenal Gland Score | | |
| Locus #1 | *CASZ1* | rs880315 | 0.96 | 4 | 0.28 | 10 | ・Enhancer chromatin in adrenal gland (active/bivalent enhancer state in multiple datasets)<br>・Open chromatin in adrenal gland (DNase/ATAC-seq peaks in multiple datasets) |
| Locus #2 | *WNT2B* | rs10776752 | 0.50 | 1f | 0.29 | 8 | ・Enhancer chromatin in adrenal gland (bivalent enhancer state in one dataset)<br>・Open chromatin in adrenal gland (ATAC-seq peaks in multiple datasets) |
| Locus #3 | *KCNK3* | rs1731243 | 0.06 | 1b | 0.61 | 10 | ・Enhancer chromatin in adrenal gland (active/weak enhancer state in multiple datasets)<br>・ZNF682 motif with TF footprint in adrenal gland |
| Locus #7 | *FBN2* | rs1004965 | 0.40 | 1b | 0.29 | 6 | ・SOX family motifs (SOX18/5/6/8) with TF footprints in non-adrenal tissues<br>・Adrenal gland eQTL for FBN2 |
| Locus #11 | *RPL23AP53* | rs11776856 | 0.30 | 1a | 0.30 | 8 | ・CTCF binding in adrenal gland (ChIP-seq peak in one dataset)<br>・Open chromatin in adrenal gland (DNase/ATAC-seq peaks in multiple datasets)<br>・CTCF motif with TF footprint in adrenal gland<br>・Adrenal gland eQTL for RP5-855D21.1, RP5-855D21.2, and RPL23AP53 |
| Locus #13 | *SLK* | rs9420907 | 0.23 | 1f | 0.32 | 8 | ・POLR2A binding in adrenal gland (ChIP-seq peak)<br>・Active promoter/enhancer chromatin in adrenal gland (active enhancer/active TSS state in multiple adrenal datasets)<br>・Open chromatin (DNase/ATAC-seq peaks in multiple datasets) |
| Locus #14 | *LSP1* | rs4980379 | 0.69 | 4 | 0.42 | 8 | ・Enhancer and promoter-proximal chromatin in adrenal gland (active/bivalent enhancer, active TSS, and flanking TSS upstream states in multiple datasets)<br>・Open chromatin in adrenal gland (DNase-seq peak in one dataset)<br>・REST motif with TF footprint in adrenal gland |

| | | | | | | | |
|---|---|---|---|---|---|---|---|
| Locus #15 | *TBX3* | rs35427 | 0.14 | 2b | 0.38 | 4 | ・POLR2A and CTCF binding in adrenal gland (ChIP-seq peaks)<br>・Enhancer chromatin in adrenal gland (active enhancer state in multiple datasets)<br>・Open chromatin in adrenal gland (DNase/ATAC-seq peaks in multiple datasets)<br>・GATA6 motif with TF footprint in adrenal gland; GATA4, LEF1, MECOM, and TCF7L2 motifs with TF footprints in non-adrenal tissues |
| Locus #16 | *RXFP2* | rs9315133 | 0.02 | 1b | 0.35 | 8 | ・CTCF binding in adrenal gland (ChIP-seq peaks)<br>・Open chromatin in adrenal gland (ATAC-seq peaks in multiple datasets)<br>・HMX2, IKZF1, IRF2, SPI1, and SPIC motifs with TF footprints in adrenal gland; multiple other motifs with TF footprints in non-adrenal tissues<br>・Adrenal gland eQTL for RXFP2 |
| Locus #18 | *ABHD17C* | rs2245049 | 0.65 | 1a | 0.40 | 10 | ・Enhancer chromatin (active enhancer state in multiple datasets)<br>・Open chromatin in adrenal gland (DNase/ATAC-seq peaks in multiple datasets)<br>・ATF2, ATF7, CREM, and JUN-family motifs with TF footprints in adrenal gland; CREB1 motif with TF footprint in non-adrenal tissues |
| Locus #19 | *MEX3C* | rs11876341 | 0.29 | 2b | 0.30 | 7 | ・Open chromatin in adrenal gland (ATAC-seq peaks in multiple datasets)<br>・NR1I3 motif with TF footprint in adrenal gland |

**Supplementary Table 8. Linkage disequilibrium metrics and Bayesian colocalization results for PA loci and adrenal gland eQTL signals.**

| Locus ID | European PA Lead SNP | Target eGene | eQTL Lead SNP | $r^2$ (PA Lead vs. eQTL Lead) | Colocalization | | | | | |
|---|---|---|---|---|---|---|---|---|---|---|
| | | | | | Analyzed Window | PP.H0 | PP.H1 | PP.H2 | PP.H3 | PP.H4 |
| Locus #3 | rs1731249 | *KCNK3* | rs1275977 | 0.79 | 2:26420025-27420025 | $2.1 \times 10^{-10}$ | $3.2 \times 10^{-5}$ | $2.9 \times 10^{-7}$ | 0.04 | 0.96 |
| | | *AC015977.6* | rs13394970 | 0.96 | | $2.4 \times 10^{-7}$ | 0.04 | $1.3 \times 10^{-7}$ | 0.02 | 0.94 |
| Locus #7 | rs13184921 | *FBN2* | rs1004965 | 0.70 | 5:127314937-128314937 | $4.3 \times 10^{-55}$ | $3.5 \times 10^{-46}$ | $1.1 \times 10^{-9}$ | 0.88 | 0.12 |
| Locus #9 | rs12190287 | *TCF21* | rs6569913 | 0.47 | 6:133695719-134695719 | $1.0 \times 10^{-22}$ | $4.3 \times 10^{-17}$ | $2.4 \times 10^{-6}$ | 0.96 | 0.04 |
| | | *TARID* | rs6934616 | 0.00 | | $9.6 \times 10^{-15}$ | $3.9 \times 10^{-9}$ | $2.4 \times 10^{-6}$ | 1.00 | $9.9 \times 10^{-6}$ |
| Locus #11 | rs59300324 | *RPL23AP53* | rs7822515 | 0.46 | 8:1-656284 | $6.4 \times 10^{-20}$ | $1.5 \times 10^{-19}$ | 0.11 | 0.27 | 0.61 |
| | | *RP5-855D21.1* | rs11774528 | 0.10 | | $3.9 \times 10^{-69}$ | $9.4 \times 10^{-69}$ | 0.29 | 0.69 | 0.02 |
| | | *RP5-855D21.2* | rs11774528 | 0.10 | | $2.7 \times 10^{-51}$ | $6.6 \times 10^{-51}$ | 0.29 | 0.69 | 0.02 |
| Locus #12 | rs55922628 | *LRMDA* | rs7078507 | 0.91 | 10:76710191-77710191 | $3.8 \times 10^{-54}$ | $1.2 \times 10^{-54}$ | 0.05 | 0.01 | 0.94 |
| | | *RP11-367B6.2* | rs7072873 | 0.66 | | 0.11 | 0.04 | 0.13 | 0.04 | 0.68 |
| Locus #16 | rs1535532 | *RXFP2* | rs9603367 | 0.42 | 13:31675002-32675002 | $2.6 \times 10^{-45}$ | $1.5 \times 10^{-14}$ | $1.7 \times 10^{-31}$ | 1.00 | 0.001 |
| Locus #18 | rs7177344 | *ABHD17C* | rs2245049 | 0.97 | 15:80497197-81497197 | $1.1 \times 10^{-5}$ | $1.2 \times 10^{-4}$ | $9.7 \times 10^{-4}$ | 0.01 | 0.99 |
| Locus #19 | rs8090393 | *MEX3C* | rs11082866 | 0.91 | 18:48290842-49290842 | $2.0 \times 10^{-4}$ | 0.002 | $5.4 \times 10^{-4}$ | 0.005 | 0.99 |

Note: For colocalization analyses, the European PA GWAS was assigned as the first trait (dataset1) and eQTL data as the second trait (dataset2).

**Supplementary Table 9. Linkage disequilibrium metrics and Bayesian colocalization results for PA loci and adrenal gland sQTL signals.**

| Locus ID | European PA Lead SNP | Target sGene | Intron ID | sQTL Lead SNP | $r^2$ (PA Lead vs. sQTL Lead) | Colocalization | | | | | |
|---|---|---|---|---|---|---|---|---|---|---|---|
| | | | | | | Analyzed Window | PP.H0 | PP.H1 | PP.H2 | PP.H3 | PP.H4 |
| Locus #14 | rs4980379 | *LSP1* | chr11:1865262:1880087:clu_9712_+ | rs1810199 | 0.60 | 11:1388614-2388614 | $1.1 \times 10^{-11}$ | $9.1 \times 10^{-5}$ | $1.8 \times 10^{-9}$ | 0.01 | 0.99 |
| | | | chr11:1869719:1880087:clu_9712_+ | rs74047621 | 0.17 | | $5.2 \times 10^{-32}$ | $4.5 \times 10^{-25}$ | $1.2 \times 10^{-7}$ | 1.00 | $2.6 \times 10^{-6}$ |

Note: For colocalization analyses, the European PA GWAS was assigned as the first trait (dataset1) and sQTL data as the second trait (dataset2).

**Supplementary Table 10. Genome-wide significant loci in the Japanese subtype-specific GWAS for APA and BAH.**

| Subtype | Prioritized Gene | Lead SNP | Chr | Position (GRCh37) | RA/NRA | Case RAF | Control RAF | OR (95% CI) | *P*-value |
|---|---|---|---|---|---|---|---|---|---|
| APA | *TARID/TCF21* | rs36125724 | 6 | 134204821 | C/T | 0.61 | 0.46 | 1.72 (1.47–2.00) | $1.7 \times 10^{-11}$ |
| BAH | *HOTTIP* | rs2240042 | 7 | 27242617 | C/T | 0.71 | 0.60 | 1.62 (1.37–1.92) | $2.5 \times 10^{-8}$ |

**Supplementary Table 11. Genome-wide significant loci from the trans-ancestry subtype-specific meta-analyses of APA and BAH.**

| Subtype | Prioritized Gene | Lead SNP | Chr | Position (GRCh37) | RA/NRA | OR (95% CI) | *P*-value |
|---|---|---|---|---|---|---|---|
| APA | *TARID/TCF21* | rs36125724 | 6 | 134204821 | C/T | 1.59 (1.41–1.79) | $2.7 \times 10^{-14}$ |
| BAH | *CASZ1* | rs880315 | 1 | 10796866 | C/T | 1.51 (1.31–1.73) | $5.5 \times 10^{-9}$ |
| | *HOTTIP* | rs1859168 | 7 | 27242359 | C/A | 1.60 (1.36–1.87) | $8.6 \times 10^{-9}$ |

**Supplementary Table 12. Subtype-specific associations and heterogeneity analyses for lead SNPs at PA risk loci.**

| Locus ID | Prioritized Gene | Lead SNP | RA/NRA | APA | | BAH | | HetISq | HetPVal |
|---|---|---|---|---|---|---|---|---|---|
| | | | | OR (95% CI) | *P*-value | OR (95% CI) | *P*-value | | |
| Locus #1 | *CASZ1* | rs880315 | C/T | 1.35 (1.18–1.53) | $6.1 \times 10^{-6}$ | 1.51 (1.31–1.73) | $5.5 \times 10^{-9}$ | 28.1 | 0.24 |
| Locus #2 | *WNT2B* | rs10776752 | T/G | 1.28 (1.10–1.49) | 0.002 | 1.39 (1.20–1.62) | $1.8 \times 10^{-5}$ | 0 | 0.43 |
| Locus #3 | *KCNK3* | rs1731249 | T/A | 1.25 (1.08–1.43) | 0.002 | 1.31 (1.13–1.52) | $4.1 \times 10^{-4}$ | 0 | 0.64 |
| Locus #4 | *CACNA1D* | rs3821843 | A/G | 1.20 (1.06–1.37) | 0.005 | 1.34 (1.17–1.53) | $1.6 \times 10^{-5}$ | 27 | 0.24 |
| Locus #5 | *CLRN1* | rs7628902 | T/C | 1.11 (0.96–1.28) | 0.16 | 1.21 (1.05–1.41) | 0.01 | 0 | 0.39 |
| Locus #6 | *CXXC4*[a] | rs71599015 | A/G | | | – | | | |
| Locus #7 | *FBN2* | rs1004965 | C/T | 1.32 (1.16–1.51) | $2.5 \times 10^{-5}$ | 1.44 (1.26–1.65) | $8.9 \times 10^{-8}$ | 0 | 0.36 |
| Locus #8 | *RSPO3* | rs4897196 | T/A | 1.19 (1.05–1.35) | 0.007 | 1.27 (1.11–1.45) | $5.8 \times 10^{-4}$ | 0 | 0.51 |
| Locus #9 | *TARID/TCF21* | rs12192720 | G/A | 1.55 (1.37–1.76) | $1.1 \times 10^{-11}$ | 1.09 (0.95–1.25) | 0.22 | 92.7 | $2.1 \times 10^{-4}$ |
| Locus #10 | *HOTTIP* | rs3735533 | C/T | 1.35 (1.16–1.57) | $1.2 \times 10^{-4}$ | 1.58 (1.35–1.85) | $1.4 \times 10^{-8}$ | 48.6 | 0.16 |
| Locus #11 | *RPL23AP53*[a] | rs2906331 | C/T | 1.01 (0.89–1.15) | 0.85 | 1.07 (0.94–1.22) | 0.31 | 0 | 0.54 |
| Locus #12 | *LRMDA* | rs7072873 | C/T | 1.32 (1.16–1.49) | $1.1 \times 10^{-5}$ | 1.22 (1.07–1.40) | 0.003 | 0 | 0.43 |
| Locus #13 | *SLK*[a] | rs9419958 | T/C | | | – | | | |
| Locus #14 | *LSP1* | rs4980379 | T/C | 1.31 (1.16–1.49) | $1.9 \times 10^{-5}$ | 1.25 (1.09–1.43) | 0.001 | 0 | 0.60 |
| Locus #15 | *TBX3* | rs35429 | A/G | 1.23 (1.07–1.41) | 0.004 | 1.26 (1.08–1.46) | 0.003 | 0 | 0.81 |
| Locus #16 | *RXFP2* | rs9603367 | T/C | 1.33 (1.14–1.55) | $3.0 \times 10^{-4}$ | 1.51 (1.26–1.80) | $5.4 \times 10^{-6}$ | 12.4 | 0.29 |
| Locus #17 | *KLF5*[a] | rs78716205 | A/G | | | – | | | |
| Locus #18 | *ABHD17C* | rs2245049 | C/T | 1.17 (1.02–1.34) | 0.03 | 1.36 (1.18–1.56) | $1.8 \times 10^{-5}$ | 54.7 | 0.14 |
| Locus #19 | *MEX3C* | rs11082867 | C/A | 1.21 (1.05–1.41) | 0.01 | 1.12 (0.95–1.31) | 0.17 | 0 | 0.47 |

[a] At loci with missing data at the lead SNP, we used a proxy SNP where possible; for all loci except *RPL23AP53*, no proxy SNPs could be identified.

**Supplementary Table 13. SNPs and effect weights used to construct ancestry-specific PA GRS.**

| Locus ID | SNP | Chr | Position (GRCh37) | EA | Weight | | |
|---|---|---|---|---|---|---|---|
| | | | | | European GRS | Japanese GRS | Modified Japanese GRS |
| Locus #1 | rs880315 | 1 | 10796866 | C | 0.279 | 0.255 | 0.255 |
| Locus #2 | rs10776752 | 1 | 113044328 | T | 0.326 | 0.352 | 0.352 |
| Locus #3 | rs1731249 | 2 | 26920025 | A | -0.205 | -0.317 | -0.317 |
| Locus #4 | rs3821843 | 3 | 53558012 | A | 0.143 | 0.259 | 0.259 |
| Locus #5 | rs7628902 | 3 | 150675697 | C | not included | -0.202 | -0.202 |
| Locus #6 | rs71599015 | 4 | 105634811 | G | -0.510 | not included | not included |
| Locus #7 | rs1004965 | 5 | 127814937 | C | 0.224 | 0.347 | 0.347 |
| Locus #8 | rs4897196 | 6 | 127225202 | T | 0.117 | 0.222 | 0.222 |
| Locus #9 | rs12192720 | 6 | 134195719 | A | -0.235 | -0.240 | not included |
| Locus #10 | rs3735533 | 7 | 27245893 | C | 0.337 | 0.402 | 0.402 |
| Locus #11 | rs71512836 | 8 | 156284 | A | 0.161 | not included | not included |
| Locus #12 | rs7072873 | 10 | 77210191 | T | -0.122 | -0.189 | -0.189 |
| Locus #13 | rs9419958 | 10 | 105675946 | C | -0.229 | not included | not included |
| Locus #14 | rs4980379 | 11 | 1888614 | T | 0.233 | 0.166 | 0.166 |
| Locus #15 | rs35429 | 12 | 115555867 | G | -0.180 | -0.204 | -0.204 |
| Locus #16 | rs9603367 | 13 | 32175002 | T | 0.363 | not included | not included |
| Locus #17 | rs78716205 | 13 | 73832307 | A | 0.420 | not included | not included |
| Locus #18 | rs2245049 | 15 | 80997197 | C | 0.168 | 0.185 | 0.185 |
| Locus #19 | rs11082867 | 18 | 48790842 | C | 0.164 | not included | not included |

**Supplementary Table 14. Association of modified GRS with biochemical markers of aldosterone excess among Japanese PA cases.**

| Parameter | Analysis | BAH (*n* = 294) | | | | All APA (*n* = 301) | | | | *KCNJ5*+ APA (*n* = 78) | | | |
|---|---|---|---|---|---|---|---|---|---|---|---|---|---|
| | | Available *n* | β | SE | *P*-value | Available *n* | β | SE | *P*-value | Available *n* | β | SE | *P*-value |
| PRA (ng/mL/h, log-transformed) | Unadjusted | 291 | 0.006 | 0.017 | 0.71 | 294 | -0.027 | 0.016 | 0.09 | 78 | -0.052 | 0.028 | 0.07 |
| | Adjusted for age and sex | | 0.003 | 0.017 | 0.85 | | -0.028 | 0.016 | 0.08 | | -0.056 | 0.029 | 0.06 |
| PAC (ng/dL, log-transformed) | Unadjusted | 292 | 0.021 | 0.011 | 0.06 | 295 | -0.004 | 0.016 | 0.83 | 78 | 0.009 | 0.033 | 0.78 |
| | Adjusted for age and sex | | 0.019 | 0.011 | 0.10 | | -0.007 | 0.016 | 0.67 | | 0.002 | 0.034 | 0.96 |
| ARR (ng/dL per ng/mL/h, log-transformed) | Unadjusted | 290 | 0.011 | 0.018 | 0.55 | 294 | 0.016 | 0.024 | 0.51 | 78 | 0.062 | 0.048 | 0.20 |
| | Adjusted for age and sex | | 0.010 | 0.018 | 0.58 | | 0.013 | 0.024 | 0.59 | | 0.058 | 0.049 | 0.24 |
| U-Ald (μg/24h, log-transformed) | Unadjusted | 226 | 0.058 | 0.018 | 0.001 | 243 | 0.019 | 0.020 | 0.36 | 67 | 0.022 | 0.037 | 0.55 |
| | Adjusted for age and sex | | 0.050 | 0.017 | 0.004 | | 0.012 | 0.020 | 0.54 | | 0.004 | 0.036 | 0.90 |
| Serum potassium (mmol/L) | Unadjusted | 294 | -0.053 | 0.022 | 0.02 | 296 | -0.002 | 0.029 | 0.94 | 78 | 0.001 | 0.063 | 0.99 |
| | Adjusted for age and sex | | -0.050 | 0.022 | 0.03 | | 0.000 | 0.029 | 0.99 | | 0.004 | 0.065 | 0.96 |

**Supplementary Table 15. Genetic correlations among PA, systolic blood pressure, and cardiovascular disease outcomes.**

| Trait 1 | Trait 2 | European | | | Japanese | | |
|---|---|---|---|---|---|---|---|
| | | $r_g$ | SE | *P*-value | $r_g$ | SE | *P*-value |
| PA | SBP | 0.46 | 0.06 | $2.3 \times 10^{-13}$ | 0.54 | 0.11 | $9.0 \times 10^{-7}$ |
| PA | CAD | 0.23 | 0.06 | $3.8 \times 10^{-5}$ | 0.24 | 0.11 | 0.02 |
| PA | AF | 0.21 | 0.06 | $5.0 \times 10^{-4}$ | 0.15 | 0.13 | 0.25 |
| PA | HF | 0.38 | 0.06 | $8.0 \times 10^{-10}$ | 0.48 | 0.19 | 0.01 |
| PA | IS | 0.44 | 0.07 | $2.1 \times 10^{-9}$ | 0.41 | 0.16 | 0.01 |
| SBP | CAD | 0.34 | 0.02 | $8.0 \times 10^{-52}$ | 0.34 | 0.08 | $1.1 \times 10^{-5}$ |
| SBP | AF | 0.14 | 0.03 | $3.3 \times 10^{-7}$ | 0.13 | 0.06 | 0.02 |
| SBP | HF | 0.37 | 0.02 | $2.9 \times 10^{-58}$ | 0.52 | 0.07 | $4.3 \times 10^{-15}$ |
| SBP | IS | 0.30 | 0.03 | $4.4 \times 10^{-18}$ | 0.45 | 0.07 | $2.2 \times 10^{-9}$ |
| CAD | AF | 0.21 | 0.02 | $1.3 \times 10^{-17}$ | -0.12 | 0.09 | 0.19 |
| CAD | HF | 0.65 | 0.02 | $5.4 \times 10^{-220}$ | 0.28 | 0.10 | 0.0031 |
| CAD | IS | 0.51 | 0.03 | $2.7 \times 10^{-53}$ | 0.19 | 0.12 | 0.10 |
| AF | HF | 0.50 | 0.03 | $4.9 \times 10^{-76}$ | 0.62 | 0.07 | $7.7 \times 10^{-19}$ |
| AF | IS | 0.40 | 0.04 | $2.6 \times 10^{-27}$ | 0.16 | 0.09 | 0.08 |
| HF | IS | 0.52 | 0.04 | $4.9 \times 10^{-48}$ | 0.49 | 0.13 | $1.0 \times 10^{-4}$ |

**Supplementary Table 16. Structural equation modeling for SBP-mediated and SBP-independent genetic effects of PA on cardiovascular disease outcomes.**

| Population | Outcome | PA → SBP (*a*) | | | SBP → Outcome (*b*) | | | PA → Outcome (*c′*) | | | SBP-independent Proportion | | |
|---|---|---|---|---|---|---|---|---|---|---|---|---|---|
| | | Estimate | SE | *P*-value | Estimate | SE | *P*-value | Estimate | SE | *P*-value | Estimate | SE | *P*-value |
| European | CAD | 0.48 | 0.10 | $4.1 \times 10^{-6}$ | 0.30 | 0.05 | $2.5 \times 10^{-10}$ | 0.11 | 0.08 | 0.16 | 0.43 | 0.20 | 0.03 |
| | AF | 0.48 | 0.10 | $4.1 \times 10^{-6}$ | 0.07 | 0.05 | 0.19 | 0.17 | 0.09 | 0.05 | 0.84 | 0.16 | $6.1 \times 10^{-8}$ |
| | HF | 0.48 | 0.10 | $4.1 \times 10^{-6}$ | 0.23 | 0.05 | $2.9 \times 10^{-5}$ | 0.30 | 0.10 | 0.003 | 0.73 | 0.09 | $7.0 \times 10^{-16}$ |
| | IS | 0.48 | 0.10 | $4.1 \times 10^{-6}$ | 0.10 | 0.07 | 0.16 | 0.41 | 0.13 | 0.002 | 0.89 | 0.09 | $5.7 \times 10^{-25}$ |
| Japanese | CAD | 0.55 | 0.20 | 0.006 | 0.31 | 0.11 | 0.005 | 0.08 | 0.15 | 0.61 | 0.31 | 0.48 | 0.51 |
| | AF | 0.55 | 0.20 | 0.006 | 0.04 | 0.14 | 0.78 | 0.15 | 0.21 | 0.49 | 0.87 | 0.53 | 0.10 |
| | HF | 0.55 | 0.20 | 0.006 | 0.33 | 0.19 | 0.08 | 0.31 | 0.30 | 0.29 | 0.63 | 0.31 | 0.04 |
| | IS | 0.55 | 0.20 | 0.006 | 0.31 | 0.15 | 0.03 | 0.23 | 0.23 | 0.31 | 0.57 | 0.32 | 0.07 |

**Supplementary Table 17. Baseline and confounder-adjusted structural equation models for the genetic effects of PA on cardiovascular disease outcomes.**

| Outcome | Path / Metric | Baseline Model | | | Confounder-adjusted Model | | |
|---|---|---|---|---|---|---|---|
| | | Estimate | SE | *P*-value | Estimate | SE | *P*-value |
| CAD | PA → SBP (*a*) | 0.48 | 0.10 | $4.1 \times 10^{-6}$ | 0.46 | 0.11 | $1.5 \times 10^{-5}$ |
| | SBP → Outcome (*b*) | 0.30 | 0.05 | $2.5 \times 10^{-10}$ | 0.26 | 0.05 | $6.8 \times 10^{-9}$ |
| | PA → Outcome (*c'*) | 0.11 | 0.08 | 0.16 | 0.05 | 0.07 | 0.54 |
| | SBP-independent Proportion | 0.43 | 0.20 | 0.03 | 0.27 | 0.35 | 0.43 |
| AF | PA → SBP (*a*) | 0.48 | 0.10 | $4.1 \times 10^{-6}$ | 0.46 | 0.11 | $1.5 \times 10^{-5}$ |
| | SBP → Outcome (*b*) | 0.07 | 0.05 | 0.19 | 0.06 | 0.05 | 0.20 |
| | PA → Outcome (*c'*) | 0.17 | 0.09 | 0.05 | 0.14 | 0.08 | 0.11 |
| | SBP-independent Proportion | 0.84 | 0.16 | $6.1 \times 10^{-8}$ | 0.83 | 0.18 | $5.9 \times 10^{-6}$ |
| HF | PA → SBP (*a*) | 0.48 | 0.10 | $4.1 \times 10^{-6}$ | 0.46 | 0.11 | $1.5 \times 10^{-5}$ |
| | SBP → Outcome (*b*) | 0.23 | 0.05 | $2.9 \times 10^{-5}$ | 0.19 | 0.05 | $5.3 \times 10^{-5}$ |
| | PA → Outcome (*c'*) | 0.30 | 0.10 | 0.003 | 0.20 | 0.09 | 0.02 |
| | SBP-independent Proportion | 0.73 | 0.09 | $7.0 \times 10^{-16}$ | 0.70 | 0.12 | $1.8 \times 10^{-8}$ |
| IS | PA → SBP (*a*) | 0.48 | 0.10 | $4.1 \times 10^{-6}$ | 0.46 | 0.11 | $1.5 \times 10^{-5}$ |
| | SBP → Outcome (*b*) | 0.10 | 0.07 | 0.16 | 0.08 | 0.07 | 0.28 |
| | PA → Outcome (*c'*) | 0.41 | 0.13 | 0.002 | 0.38 | 0.13 | $4.3 \times 10^{-3}$ |
| | SBP-independent Proportion | 0.89 | 0.09 | $5.7 \times 10^{-25}$ | 0.91 | 0.09 | $4.1 \times 10^{-25}$ |

**Supplementary Table 18. Association of lead SNPs at PA risk loci with SBP and serum potassium in Europeans.**

| Locus ID | Prioritized Gene | Lead SNP | Chr | Position (GRCh37) | PA RA/NRA | PA | | | SBP | | | Serum Potassium | | |
|---|---|---|---|---|---|---|---|---|---|---|---|---|---|---|
| | | | | | | β | SE | *P*-value | β | SE | *P*-value | β | SE | *P*-value |
| Locus #1 | *CASZ1* | rs880315 | 1 | 10796866 | C/T | 0.279 | 0.031 | $2.5 \times 10^{-19}$ | 0.454 | 0.026 | $6.7 \times 10^{-71}$ | -0.041 | 0.003 | $2.7 \times 10^{-59}$ |
| Locus #2 | *WNT2B* | rs10776752 | 1 | 113044328 | T/G | 0.326 | 0.045 | $3.5 \times 10^{-13}$ | 0.725 | 0.046 | $7.3 \times 10^{-56}$ | -0.050 | 0.005 | $1.6 \times 10^{-25}$ |
| Locus #3 | *KCNK3* | rs1731249 | 2 | 26920025 | T/A | 0.205 | 0.031 | $2.9 \times 10^{-11}$ | 0.518 | 0.025 | $1.3 \times 10^{-97}$ | -0.036 | 0.003 | $7.7 \times 10^{-46}$ |
| Locus #4 | *CACNA1D* | rs3821843 | 3 | 53558012 | A/G | 0.143 | 0.034 | $3.0 \times 10^{-5}$ | 0.307 | 0.027 | $6.3 \times 10^{-30}$ | -0.021 | 0.003 | $4.3 \times 10^{-14}$ |
| Locus #6 | *CXXC4* | rs71599015 | 4 | 105634811 | A/G | 0.510 | 0.093 | $4.6 \times 10^{-8}$ | | – | | -0.004 | 0.007 | 0.56 |
| Locus #7 | *FBN2* | rs1004965 | 5 | 127814937 | C/T | 0.224 | 0.033 | $8.3 \times 10^{-12}$ | 0.282 | 0.026 | $6.8 \times 10^{-28}$ | -0.032 | 0.003 | $1.1 \times 10^{-33}$ |
| Locus #8 | *RSPO3* | rs4897196 | 6 | 127225202 | T/A | 0.117 | 0.031 | $2.0 \times 10^{-4}$ | 0.367 | 0.025 | $2.9 \times 10^{-50}$ | -0.028 | 0.002 | $6.5 \times 10^{-30}$ |
| Locus #9 | *TARID/TCF21* | rs12192720 | 6 | 134195719 | G/A | 0.235 | 0.037 | $1.4 \times 10^{-10}$ | 0.181 | 0.027 | $1.7 \times 10^{-11}$ | -0.029 | 0.003 | $2.8 \times 10^{-24}$ |
| Locus #10 | *HOTTIP* | rs3735533 | 7 | 27245893 | C/T | 0.337 | 0.061 | $2.8 \times 10^{-8}$ | 0.809 | 0.046 | $1.1 \times 10^{-69}$ | -0.072 | 0.005 | $4.2 \times 10^{-54}$ |
| Locus #11 | *RPL23AP53* | rs71512836 | 8 | 156284 | A/C | 0.161 | 0.038 | $1.8 \times 10^{-5}$ | 0.170 | 0.034 | $6.7 \times 10^{-7}$ | -0.019 | 0.003 | $6.1 \times 10^{-11}$ |
| Locus #12 | *LRMDA* | rs7072873 | 10 | 77210191 | C/T | 0.122 | 0.031 | $7.8 \times 10^{-5}$ | 0.013 | 0.024 | 0.60 | -0.003 | 0.003 | 0.23 |
| Locus #13 | *SLK* | rs9419958 | 10 | 105675946 | T/C | 0.229 | 0.044 | $2.2 \times 10^{-7}$ | 0.265 | 0.042 | $2.6 \times 10^{-10}$ | -0.029 | 0.004 | $2.5 \times 10^{-15}$ |
| Locus #14 | *LSP1* | rs4980379 | 11 | 1888614 | T/C | 0.233 | 0.031 | $6.7 \times 10^{-14}$ | 0.491 | 0.025 | $4.1 \times 10^{-83}$ | -0.045 | 0.003 | $1.1 \times 10^{-67}$ |
| Locus #15 | *TBX3* | rs35429 | 12 | 115555867 | A/G | 0.180 | 0.033 | $3.3 \times 10^{-8}$ | 0.391 | 0.025 | $2.1 \times 10^{-55}$ | -0.040 | 0.003 | $2.5 \times 10^{-54}$ |
| Locus #16 | *RXFP2* | rs9603367 | 13 | 32175002 | T/C | 0.363 | 0.031 | $2.9 \times 10^{-31}$ | 0.269 | 0.025 | $7.9 \times 10^{-28}$ | -0.054 | 0.003 | $1.9 \times 10^{-100}$ |
| Locus #17 | *KLF5* | rs78716205 | 13 | 73832307 | A/G | 0.420 | 0.066 | $2.5 \times 10^{-10}$ | 0.468 | 0.059 | $2.7 \times 10^{-15}$ | -0.067 | 0.006 | $1.1 \times 10^{-28}$ |
| Locus #18 | *ABHD17C* | rs2245049 | 15 | 80997197 | C/T | 0.168 | 0.035 | $1.8 \times 10^{-6}$ | 0.241 | 0.027 | $1.1 \times 10^{-18}$ | -0.024 | 0.003 | $2.3 \times 10^{-17}$ |
| Locus #19 | *MEX3C* | rs11082867 | 18 | 48790842 | C/A | 0.164 | 0.033 | $8.9 \times 10^{-7}$ | 0.217 | 0.027 | $3.6 \times 10^{-16}$ | -0.025 | 0.003 | $7.1 \times 10^{-21}$ |

**Supplementary Table 19. Association of PA GRS with SBP, serum potassium, and PAC in population-based cohorts.**

| Target Cohort | Target Trait | *n* | β | SE | *P*-value |
|---|---|---|---|---|---|
| BBJ | SBP (mmHg) | 35,160 | 0.55 | 0.09 | $1.1 \times 10^{-10}$ |
| | Serum potassium (mmol/L) | 46,724 | -0.013 | 0.001 | $< 2.2 \times 10^{-16}$ |
| KING | SBP (mmHg) | 2,453 | 0.99 | 0.41 | 0.01 |
| AoU | SBP (mmHg) | 114,093 | 0.46 | 0.04 | $< 2.2 \times 10^{-16}$ |
| | Serum potassium (mmol/L) | 129,856 | -0.013 | 0.001 | $< 2.2 \times 10^{-16}$ |
| | PAC (ng/dL, log-transformed) | 838 | 0.044 | 0.016 | 0.005 |

**Supplementary Table 20. Overlap in causative/responsible genes among variants of different PA pathologies.**

| Gene Locus | Familial PA | Sporadic PA (Present GWAS) | | | APA Somatic Mutations |
|---|---|---|---|---|---|
| | Causative Gene | Lead SNP | *P*-value | Nearby Gene | Causative Gene |
| *CYP11B2* | *CYP11B1/CYP11B2* | rs3750247[a] | $1.4 \times 10^{-7}$ | *CYP11B1/CYP11B2* | — |
| *CLCN2* | *CLCN2* | | — | | *CLCN2* |
| *KCNJ5* | *KCNJ5* | | — | | *KCNJ5* |
| *CACNA1H* | *CACNA1H* | | — | | *CACNA1H* |
| *CACNA1D* | — | rs3821843 | $1.8 \times 10^{-10}$ | *CACNA1D* | *CACNA1D* |
| *ATP1A1* | — | | — | | *ATP1A1* |
| *ATP2B3* | — | | — | | *ATP2B3* |

[a] A suggestive association between rs145725189 and PA at the *CYP11B1/CYP11B2* locus has been previously reported[4]. Although this same SNP was not included in the PA association analysis of the present GWAS, a borderline association was also observed for rs3750247, which is located in the vicinity of rs145725189.

### Supplementary Methods

#### Study Cohorts with Genotype Data

- **Japanese PA cohort**: Patients with primary aldosteronism (PA) in the Japanese PA Study (JPS) cohort were recruited from three medical centers and a biobank network in Japan: Tohoku University Hospital, Yokohama Rosai Hospital, Institute of Science Tokyo Hospital, and the National Center Biobank Network (NCBN).
- **BBJ cohort**: BioBank Japan (BBJ) is a large-scale clinical biobank in Japan that was launched in 2003. Through partnerships with 12 medical institutions nationwide, DNA and serum samples have been collected from approximately 270,000 patients diagnosed with 51 target diseases. Details of the genotyping and quality control procedures have been described previously[5,6]. Among the combined 1$^{st}$ and 2$^{nd}$ BBJ cohorts with available genotype data (accession JGAS000412; *n* = 54,405), individuals were classified as non-hypertensive for inclusion in the control group of the Japanese PA genome-wide association study (GWAS) if they met all of the following criteria: (1) no history of essential hypertension; (2) systolic blood pressure (SBP) ≤ 140 mmHg; (3) diastolic blood pressure ≤ 90 mmHg; and (4) no use of antihypertensive medication.
- **The Kita-Nagoya Genomic Epidemiology (KING) study:** The KING study (ClinicalTrials.gov identifier: NCT00262691) is an ongoing community-based prospective observational study of the genetic basis of cardiovascular disease and its risk factors. The study recruited 3,975 Japanese subjects aged 50-80 years, who underwent community-based annual health checkups between May 2005 and December 2007. A subset of the KING study sample (*n* = 2,453) was included in the PA genetic risk score (GRS) analysis; 52.0% were women and the average age was 64.1 years. As previously reported[7], the participants were genotyped using the Illumina 660-Quad, Omni2, OmniExpress-12, and/or OmniExpress-24 arrays. Quality control and imputation were performed separately for each array. The 1000 Genomes Project (1kGP)[8] Phase 3 all-ancestry reference panel was used for imputation. PA GRSs were calculated for each individual; for the 191 participants genotyped on multiple arrays, the final PA GRS was calculated as the average of their array-specific GRSs. Blood pressure was measured using the USM-700G machine (Elquest Co., Japan) in the seated position, and the average of two readings was used for analysis.
- **The All of Us (AoU) Research Program cohort**: The AoU program is a large-scale U.S. precision medicine initiative launched by the National Institutes of Health in 2018.

It includes a diverse cohort of approximately 630,000 U.S. participants enrolled at age 18 years or older, linking survey data, electronic health records, physical measurements, and genomic information. In this study, analyses were restricted to participants of European ancestry with available clinical data for each trait. The number of participants for each analysis is provided in Section: *GRS Calculation and Exclusion Criteria*.

**Classification of PA Subtypes**

Depending on the diagnostic modalities available at each enrollment site, PA patients were classified into subtypes—aldosterone-producing adenoma (APA), bilateral adrenal hyperplasia (BAH), or inconclusive—based on adrenal venous sampling (AVS), super-selective AVS (ss-AVS), clinical evaluation, and/or a validated prediction score.

Among 282 cases from Tohoku University Hospital and the Institute of Science Tokyo Hospital, AVS provided definitive lateralization results in 270 cases, while 12 cases without AVS or with ambiguous AVS findings were classified as inconclusive. For this study, among the 270 patients with definitive AVS results, 137 with unilateral aldosterone secretion (unilateral PA) were classified as APA, and 133 with bilateral aldosterone secretion (bilateral PA) as BAH. This AVS-based classification was used regardless of subsequent histopathological subtype. Among the unilateral PA cases, 87.5% (120/137) had an adrenal nodule or mass on computed tomography and/or magnetic resonance imaging that corresponded to the side of aldosterone lateralization on AVS. Post-operative histopathological assessment with CYP11B2 immunohistochemistry was available for 92 resected specimens and classified according to the international histopathology consensus for unilateral PA[9]: 93.5% (86/92) showed classical APA, and 6.5% (6/92) showed non-classical lesions (e.g., MAPN/MAPM).

At Yokohama Rosai Hospital, ss-AVS enabled segment-level assessment of aldosterone secretion. Of 262 patients evaluated in this manner, 125 cases with suspected adenomatous lesions were classified as APA, regardless of whether the lesions were unilateral or bilateral; 102 cases with suspected bilateral hyperplastic changes were classified as BAH; and 35 cases with suspected mixed pathology (e.g., concurrent adenoma and hyperplasia) were classified as inconclusive.

Among 126 patients from the NCBN, APA was assigned according to the attending endocrinologist's diagnosis, based on available clinical findings. Thirty-nine patients were classified as having APA, of whom 33 had undergone AVS. Among the remaining 87 patients not diagnosed with APA, a validated clinical prediction score[10] was applied: 59 patients with scores ≥ 8 were classified as BAH, and 28 patients with scores < 8 as inconclusive.

Thus, across the entire cohort, 301 patients were classified as having APA, 294 as having BAH, and 75 as inconclusive.

#### Somatic Mutation Profiling in APA Tumors

Somatic mutation status was assessed in 97 of 301 APA cases for which tumor tissue was available. Genomic DNA was analyzed stepwise, beginning with Sanger sequencing of hotspot regions in *KCNJ5*. Samples without *KCNJ5* variants were subsequently screened for other established driver genes, including *CACNA1D*, *ATP1A1*, *ATP2B3*, and *CTNNB1*, using targeted next-generation sequencing or Sanger sequencing. This approach identified somatic mutations in 91 cases: *KCNJ5* ($n$ = 78), *CACNA1D* ($n$ = 6), *ATP2B3* ($n$ = 4), *ATP1A1* ($n$ = 2), and *CTNNB1* ($n$ = 1). The remaining six sequenced samples were classified as wild-type with respect to somatic mutations in known driver genes.

#### Biochemical Markers of Aldosterone Excess

Hormonal and electrolyte markers of aldosterone excess were collected at diagnosis, including plasma renin activity (PRA, ng/mL/h), plasma aldosterone concentration (PAC, ng/dL), aldosterone-to-renin ratio (ARR, ng/dL per ng/mL/h), 24-hour urinary aldosterone excretion (U-Ald, μg/24h), and serum potassium (mmol/L). In patients receiving potassium supplementation, serum potassium was set to the lower of 3.5 mmol/L or the measured value. PAC and U-Ald were measured using radioimmunoassay (RIA) or chemiluminescent enzyme immunoassay (CLEIA), with CLEIA-derived values converted to RIA-equivalent values as previously described[11,12]. U-Ald measurements were performed under a controlled sodium intake of 10–12 g/day.

#### Genotype Data Processing, Quality Control, and Imputation

Genomic DNA from recruited PA patients was genotyped on the Illumina Infinium Asian Screening Array. BBJ genotype data were obtained from the National Bioscience Database Center (accession JGAS000412). Raw IDAT files were processed and aligned to the human reference genome (GRCh37) using bcftools with the idat2gtc and gtc2vcf plugins (https://software.broadinstitute.org/software/gtc2vcf/). Variants with poor clustering (GenTrain score < 0.8 or cluster separation < 0.6), minor allele frequency

(MAF) < 0.005, call rate < 0.95, or Hardy–Weinberg equilibrium $P < 1.0 \times 10^{-6}$ were excluded. Samples were removed if they had a low call rate (< 0.95), sex discordance, or cryptic relatedness (KING kinship > 0.0884). Additionally, we inferred ancestry by principal component analysis (PCA) with HapMap3[13] reference panels and retained only individuals of Japanese ancestry.

Phasing was performed with SHAPEIT[14] using a standard genetic map, and imputation with Minimac3[15] using the 1kGP Phase 3 reference panel. Post-imputation, we retained only autosomal, biallelic single nucleotide polymorphisms (SNPs) meeting MAF-stratified imputation quality filters (imputation $r^2 \geq 0.8$ for 0.005-0.01, ≥ 0.6 for 0.01-0.05, and ≥ 0.3 for ≥ 0.05) for association analyses; 6,646,529 autosomal biallelic SNPs were analyzed.

**External PA GWAS Datasets for Trans-Ancestry Meta-analysis**

To perform a trans-ancestry GWAS meta-analysis of PA, we obtained publicly available summary statistics from large biobanks and a published cohort resource, as detailed below.

**FinnGen–MVP–UKBB resource**: The FinnGen–MVP–UKBB meta-analysis resource (https://public-mvp-ukbb.finngen.fi/) is a central repository that offers GWAS summary statistics from multiple large-scale biobanks and their meta-analyzed results. This platform employs a harmonized endpoint-mapping framework based primarily on ICD-10 code overlap and curated phenotype concordance to enable cross-cohort and trans-ancestry meta-analyses. For the present study, we used summary statistics corresponding to hyperaldosteronism (ICD-10: E26; FinnGen endpoint: E4_hyperaldosteronism). Detailed descriptions of the resource and its data processing procedures are available in the FinnGen public documentation (https://storage.googleapis.com/finngen-public-data-r12/meta_analysis/mvp_ukbb/README). A brief overview of each contributing cohort and its ancestral composition is provided below.

- **FinnGen**[16]: FinnGen is a large-scale genomics initiative initiated in 2017 that has analyzed data from over 500,000 Finnish biobank participants, linking genetic information with national health registries to investigate disease mechanisms and genetic predisposition. Participants are predominantly of Finnish ancestry. For this study, we used data from FinnGen Data Freeze 12 (R12).
- **UK Biobank (UKBB)**[17]: UK Biobank is a population-based prospective cohort study in the United Kingdom comprising ~500,000 participants aged 40-69 at recruitment

(2006-2010). In the FinnGen–MVP–UKBB meta-analysis resource, only the European-ancestry subset is included.

- **Million Veteran Program (MVP)**[18]: MVP is a large-scale biobank established by the U.S. Department of Veterans Affairs in 2011 and has enrolled over 900,000 U.S. veterans representing diverse ancestral backgrounds, including European, African, and admixed American populations. For the E26 endpoint, ancestry-specific summary statistics were only available for European and African populations.

**COMETE network cohort**[19]: PA cases were recruited from the COMETE (COrtico- et MEdullo-surrénale, les Tumeurs Endocrines) network, while controls were drawn from the population-based Paris Prospective Study III (PPS3) cohort. Diagnosis of PA followed institutional protocols consistent with the Endocrine Society clinical practice guidelines. Diagnostic evaluation included screening with the plasma ARR, confirmatory suppression testing to demonstrate autonomous aldosterone secretion, and adrenal imaging for structural assessment. The discovery dataset comprised 562 individuals with PA and 950 controls, all of European ancestry. Subtype classification was available for 561 of 562 PA cases; 321 were diagnosed with unilateral APA and 240 with BAH based on CT, MRI, and/or AVS.

**MR-MEGA Analysis**

Meta-regression of multi-ethnic genetic association studies (MR-MEGA)[20] was used to conduct an ancestry-aware meta-analysis and to partition effect-size heterogeneity across cohorts into ancestry-correlated and residual components. As allele-frequency data were unavailable for the COMETE cohort, frequencies from the 1kGP European reference panel were substituted, following recommendations from the original publication[20]. Ancestry adjustment was performed using two principal components (PCs) (--pc 2), as the first two axes of genetic variation (PC0 and PC1) clearly separated the three ancestry clusters (Supplementary Fig. 15).

**Locus-Specific Conditional and Heterogeneity Analyses at the *TARID/TCF21* Locus**

For locus-specific follow-up analyses at the *TARID/TCF21* locus, we performed conditional analyses to dissect subtype-specific association patterns across ancestries. In European datasets, analyses were conducted using GCTA-COJO[21,22] with linkage disequilibrium estimated from the 1kGP European reference panel, conditioning on representative SNPs (PA: rs12190287; APA: rs36125724; BAH: rs1336170). In the Japanese cohort, conditional analyses were performed using REGENIE[23] with individual-level genotype data under the same framework as the primary and subtype-stratified GWAS, with genotype dosages of the corresponding lead variants included as covariates in the model, either individually or jointly.

**Selection of European SBP Datasets**

For cross-trait analyses in Europeans, we used two distinct SBP datasets tailored to different analytical goals. For locus-level comparisons at PA lead SNPs, evaluating the concordance of their effects on PA, SBP, and serum potassium, we used body mass index (BMI)-adjusted SBP summary statistics from a large GWAS meta-analysis (GCST90310294) to maximize statistical power, acknowledging that this may introduce covariate-induced bias at specific loci. For genome-wide analyses, including LD score regression (LDSC)[24] and the latent causal variable (LCV)[25] model, we used the smaller Pan-UK Biobank dataset without BMI adjustment (GCST90691848) to avoid genome-wide bias.

**GRS Calculation and Exclusion Criteria**

GRSs were calculated separately for each cohort using PLINK[26] with the --score function, which computes a weighted sum of risk alleles at the specified SNPs. In the BBJ cohort, GRSs were evaluated in the combined first and second cohorts (*n* = 51,071) to characterize overall score distributions; this includes, but is not limited to, the non-hypertensive BBJ subset (*n* = 25,998) used in the case-control genome-wide association analysis (see Section: *Study Cohorts with Genotype Data*). The KING study cohort was used as the Japanese reference for score standardization, as BBJ is a disease-based biobank and is therefore less representative of the general population distribution. For European analyses, GRSs were evaluated in the European subset of the AoU cohort, using data from the All of Us Research Program Controlled Tier Dataset version 8, available to authorized users on the Research Workbench (workspace aou-rw-f61c46ea).

In population-based cohorts, associations between GRSs and quantitative traits were evaluated. Data availability differed by trait: SBP was available in BBJ, KING, and AoU; serum potassium in BBJ and AoU; and PAC in AoU only. Trait-specific exclusions and resulting sample sizes for each analysis were as follows. For SBP, individuals taking antihypertensive or diuretic drugs (BBJ Drug Classification numbers 213 and 214; AoU ATC codes C02, C03, C07, C08, and C09) were excluded in BBJ and AoU, resulting in final sample sizes of 35,160 and 114,093, respectively. The KING cohort included 2,453 participants, with no medication-based exclusions applied. For serum potassium, 46,724 individuals from BBJ and 129,856 from AoU were included, with no medication-based exclusions applied. For PAC, participants taking aldosterone antagonists or renin–angiotensin system agents (ATC codes C03DA and C09) were excluded, leaving 838 AoU participants.

For the analysis within PA cases (JPS), we constructed a modified Japanese GRS that excluded the *TARID/TCF21* locus to capture the polygenic burden without the disproportionate effect of the APA-specific signal. Using this modified GRS, we assessed associations with biochemical markers of aldosterone excess, including PRA, PAC, ARR, and U-Ald, all log-transformed, using linear regression models with and without adjustment for age and sex. Analyses were stratified by subtype (APA and BAH) and somatic mutation status (*KCNJ5*-mutated APA). In addition, the distributions of the modified GRS, U-Ald, and serum potassium were compared between APA and BAH to evaluate differences in polygenic burden and biochemical severity.
